# Gametocyte production in incident *P. falciparum* infections: a longitudinal study in a low transmission setting under intensive vector control

**DOI:** 10.1101/2022.08.29.22279332

**Authors:** Chiara Andolina, Jordache Ramjith, John Rek, Kjerstin Lanke, Joseph Okoth, Lynn Grignard, Emmanuel Arinaitwe, Jessica Briggs, Jeffrey Bailey, Ozkan Aydemir, Moses R Kamya, Bryan Greenhouse, Grant Dorsey, Sarah G Staedke, Chris Drakeley, Marianne Jonker, Teun Bousema

## Abstract

Malaria transmission depends on the presence of *Plasmodium* gametocytes that are the only parasite life stage that can infect mosquitoes. Gametocyte production varies between infections and over the course of infections. Infection duration is influenced by host and parasite characteristics, and is highly important for gametocyte production but poorly quantified. Between 2017-2019 an all-age cohort from Tororo, eastern Uganda was followed by continuous passive and routine assessments. Among 104 longitudinally monitored incident infections coming from 98 individuals, we observed that nearly all infections lasting 3 or more months initiated gametocyte production prior to clearance. However, the majority of infections were of shorter duration (<28 days) and were cleared before gametocytes were detectable. Infections in individuals with sickle-cell trait were more likely to produce gametocytes and produced gametocytes at higher densities. Our findings suggest that a large proportion of infections may be too short in duration and of too low density to contribute to onward transmission.

## Introduction

Despite significant progress in the last decade, *P. falciparum* malaria remains a leading cause of morbidity and mortality in many countries in Sub Saharan Africa. In 2020 there were 241 million malaria cases, 14 million more than in 2019, and the number of deaths increased from 558,000 to 627,000 (Who, 2021). This was in part attributed to the disruption of malaria prevention and treatment during the COVID-19 pandemic (Heuschen et al., 2021). However, even before the COVID-19 pandemic, global progress against malaria plateaued or even reversed in some endemic regions, related to funding shortages that impacted access to vector control, early diagnosis and treatment. Control efforts are further threatened by the emergence and spread of drug and insecticide resistance (Kleinschmidt et al., 2018; Uwimana et al., 2020); new interventions are therefore urgently needed to sustainably reduce the burden of malaria and move towards elimination. Transmission-blocking interventions are considered highly relevant for elimination purposes and their development and deployment depends on a thorough understanding of human to mosquito transmission.

In human hosts, *P. falciparum* parasites replicate asexually and multiply over approximately 48 hours, after which merozoites are released into the circulation where they invade new red blood cells. The majority of merozoites continue this asexual cycle, whilst a minority commit to become male or female gametocytes (Eichner et al., 2001). Gametocytes mature in human host tissues, primarily the bone marrow and spleen (Smalley et al., 1981; Aguilar et al., 2014; Joice et al., 2014), until their release into the peripheral circulation approximately 8-12 days after the initial wave of asexual parasites (Eichner et al., 2001; Reuling et al., 2018). Mature gametocytes thereby become accessible to mosquitoes taking a blood meal. Once in the mosquito midgut, gametocytes differentiate into gametes that fertilize and, following sporogonic development, ultimately lead to the presence of infective sporozoites in the mosquito salivary glands. The average lifespan of circulating mature gametocytes is 6.5 days (Smalley et al., 1977; Eichner et al., 2001; Bousema et al., 2010) but a proportion of gametocytes can persist and be infectious to mosquitoes for several weeks or even months following effective clearance of asexual parasites (Eichner et al., 2001; Shekalaghe et al., 2007; Bousema et al., 2010; Andagalu et al., 2014).

Although gametocytes are formed from their asexual progenitors and gametocyte densities are therefore associated with the preceding asexual parasite biomass, this association is relatively weak because gametocyte production may vary between infections (Barry et al., 2021) and over the course of infections (Reece et al., 2008; Johnston et al., 2013). The factors that drive gametocyte production are poorly understood. Controlled malaria infections in malaria naïve volunteers suggest that gametocyte production may be highest early after infection, although a comparison of infections during peak and low transmission seasons suggests that gametocyte production relative to the asexual parasite biomass may be higher in the low transmission season (Drakeley et al., 2006) when infections are relatively old (Andrade et al., 2020). The duration of infection is a critical determinant of gametocyte production and thereby the human infectious reservoir for malaria but is currently poorly characterised. While *P. falciparum* infections can be very short-lived, lasting only a few days (Farnert et al., 1997; Bretscher et al., 2015; Roe et al., 2022), most attention has gone to longer-lasting infections that can persist for several months or even years in the absence of treatment (Ashley et al., 2014; Briggs et al., 2020). These longer-term infections are considered the most important drivers of malaria transmission in most endemic settings and are often present at low, sometimes submicroscopic, parasite densities. It is generally assumed that all *P. falciparum* infections, including submicroscopic infections, have the potential to produce gametocytes and be infectious to mosquitoes (Bousema et al., 2014) because of low-level stochastic commitment to gametocyte production (Kafsack et al., 2014). However, there have been no accurate estimations of gametocyte production during natural infections that require repeated sampling and sensitive molecular techniques.

*P. falciparum* gametocyte production may be stimulated by various triggers, including anaemia (Ho et al., 1990; Price et al., 1999; Von Seidlein et al., 2001; Chotivanich et al., 2002; Gbotosho et al., 2011), human genetic factors (Lawaly et al., 2010; Goncalves et al., 2017), drug treatment (Buckling et al., 1999; Peatey et al., 2009), and the presence of certain *P. falciparum* genotypes (Wampfler et al., 2014). Sickle cell trait (HbAS) is a common host genetic polymorphism in eastern Uganda, with a prevalence around 30% (Gong et al., 2012; Bwayo et al., 2014; Walakira et al., 2017). HbAS is associated with protection against severe malaria disease (Ruwende et al., 1995; Price et al., 1999; Liang et al., 2020) but its impact on transmission is incompletely understood. A single longitudinal study in an area of intense malaria transmission in Uganda where gametocytes were examined by microscopy found evidence of higher gametocyte carriage among HbAS individuals (Gong et al., 2012), a finding that was supported by a cross-sectional study from Senegal (Lawaly et al., 2010). However, other studies observed no association with gametocyte carriage but suggested increased transmissibility of gametocytes from HbAS carriers (Robert et al., 1996; Gouagna et al., 2010; Gong et al., 2012; Goncalves et al., 2017).

We examined gametocyte production over the course of infections in relation to host and parasite characteristics using longitudinal data from 104 incident malaria infections in an all-age cohort in eastern Uganda (Andolina et al., 2021). Sensitive molecular assays were deployed for the quantification of total parasite biomass and gametocyte density; amplicon deep sequencing was used to genotype infections and distinguish between incident and persistent infections. Human host and infection characteristics were examined in relation to infection duration and gametocyte production.

## Results

### The majority of incident infections were asymptomatic and of short duration

Between 2017 and 2019, all 531 residents from 80 randomly selected households were enrolled into a study cohort followed for up to 24 months (Nankabirwa et al., 2020; Andolina et al., 2021). We investigated the duration of incident malaria infections among individuals who were parasite-free by qPCR on 3 previous visits; resulting in 104 infections among 98 individuals. Sixteen of these incident infections (15.4%) were symptomatic at the time of first detection, had a geometric mean of 19,817 parasites/µL (95% CI: 7,260-54,096), and received first-line treatment for uncomplicated malaria immediately after sampling. The remainder of incident infections (84.6%) were asymptomatic and had a geometric mean of 0.87 parasites/µL (95% CI: 0.34-2.21) at the time of initial detection (**Table 1**); in 13.6% (12/88) of these initially asymptomatic infections, symptoms and parasites detected by microscopy occurred at a later time-point and prompted antimalarial treatment.

**Table 1.** Characteristics of incident infections that presented with symptoms and initially asymptomatic.

|  | Incident initially<br>symptomatic infections | Incident initially<br>asymptomatic infections | p-value |
| --- | --- | --- | --- |
| **At moment of infection**<br>**detection** |  |  |  |
| **Proportion of infections in each**<br>**class** | 15.4% (16/104) | 84.6% (88/104) |  |
| **Hb genotype** | 1 missing | 2 missing |  |
| AA | 73.3% (11/15) | 67.4% (58/86) | p=0.1435 |
| SS | 24.4% (3/15) | 8.1% (7/86) |  |
| AS | 8.1% (1/15) | 24.4% (21/86) |  |
| **Geometric mean parasite**<br>**density; parasites/μL (95% CI)** | 19817.75 (7260.10, 54096.12) | 0.87 (0.34-2.21) | p<0.0001 |
| **Gametocyte prevalence** | 6.7% (1/16) | 16.5% (14/85)*** | p=0.4544 |
| **During infection** |  |  |  |
| **Geometric mean peak parasite**<br>**density; parasites/μL (95% CI)** | 19817.75 (7260.10, 54096.12) | 5.53 (1.77, 17.23) | p<0.0001 |
| **Polyclonal infection \*** | 50% (8/16) | 33.3% (17/51) | p=0.2505 |
| **Number of clones, median**<br>**(range)\*\*** | 1.5 (1–5) | 1 (1–5) | p=0.1158 |
| **Percentage receiving**<br>**treatment\*\*\*** | 100% (16/16) | 13.6% (12/88) | p<0.0001 |

\*Polyclonal infection was defined as having at least 2 parasite clones based on AMA-1 amplicon sequencing.
\*\*Number of clones was defined as the total number of clones detected by AMA-1 amplicon sequencing. Multiclonal infection and the number of parasite clones were measured across the entire duration of infection; for symptomatic incident infections this equaled the first time-point of detection since treatment was immediately provided. For asymptomatic incident infections AMA-1 sequencing was unsuccessful for 37 individuals.\*\*\* drug treatment was initiated immediately for symptomatic individuals, while for incident initially asymptomatic infections treatment occurred in 13.6% of infections during follow-up (i.e. the infection started asymptomatic but developed into a symptomatic episode). For all cells % (n/N) is provided unless stated otherwise.

In the entire study population, including individuals without incident infections, 25.5%, 129/505 had the HbAS and 8.3%% (42/505) had the HbSS genotype, which was similar to the occurrence of this haemoglobinopathy among individuals with incident infections (21.8% HbAS (22/101) and 8.9% HbSS (10/101)). Among HbAS individuals, 4.5% (1/22) of incident infections were symptomatic upon presentation compared to 30.0% (3/10) among HbSS and 15.9% (11/69) among HbAA individuals (**Table 1**, p=0.1435).

The duration of infections was estimated based on time to qPCR negativity; if an infection was undetected at a given time-point followed by a qPCR positive visit up to 16 weeks later with the same parasite clone, we considered an infection as persisting. The detection of parasite clones was based on AMA-1 amplicon sequencing. Sequencing success was associated with parasite density, with a geometric mean parasite density of 0.03 parasites/µL (95% CI: 0.02-0.05) among samples that failed (n=37) and 59.00 parasites/µL (95% CI: 17.18, 202.69, p<0.0001) that had successful genotyping results (n=67) at the initial detection of incident infections. Overall, 37.3% (25/67) of incident infections with genotyping results were polyclonal, i.e. had multiple *P. falciparum* clones detected by AMA-1 amplicon sequencing.

In our cohort, most incident infections were only detected at a single time point (**Figure 1)**, with a steep decline in survival probabilities for clearance after 4 weeks. When focusing only on those infections that were initially asymptomatic (n=88), and thus did not receive treatment when first detected, 55.7% (49/88) were detected only once and only 19.3% (17/88) of infections persisted for 12-96 weeks. All 16 initially symptomatic infections received treatment and were thus considered immediately cleared; this was illustrated by the immediately decline in survival probabilities (**Figure 1**). The geometric mean parasite density among incident asymptomatic infections that were detected only once was 0.15 parasites/µL (95% CI: 0.06-0.38), which was lower than the initial parasite density of infections that were detected at multiple occasions (2.11 parasites/µL (95% CI: 0.62-3.59, p<0.0001). Among incident infections that were initially asymptomatic, there was no apparent association between age and infection duration: 65% (13/20), 51.4% (19/37) and 54.8% (17/31) of infections were detected only once for individuals aged <5, 5-15 and 16+ years old, respectively (**Figure S1)**. Similarly, sickle cell trait (HbAS) did not have a notable effect on infection duration (**Figure S1**).

**Figure 1.**
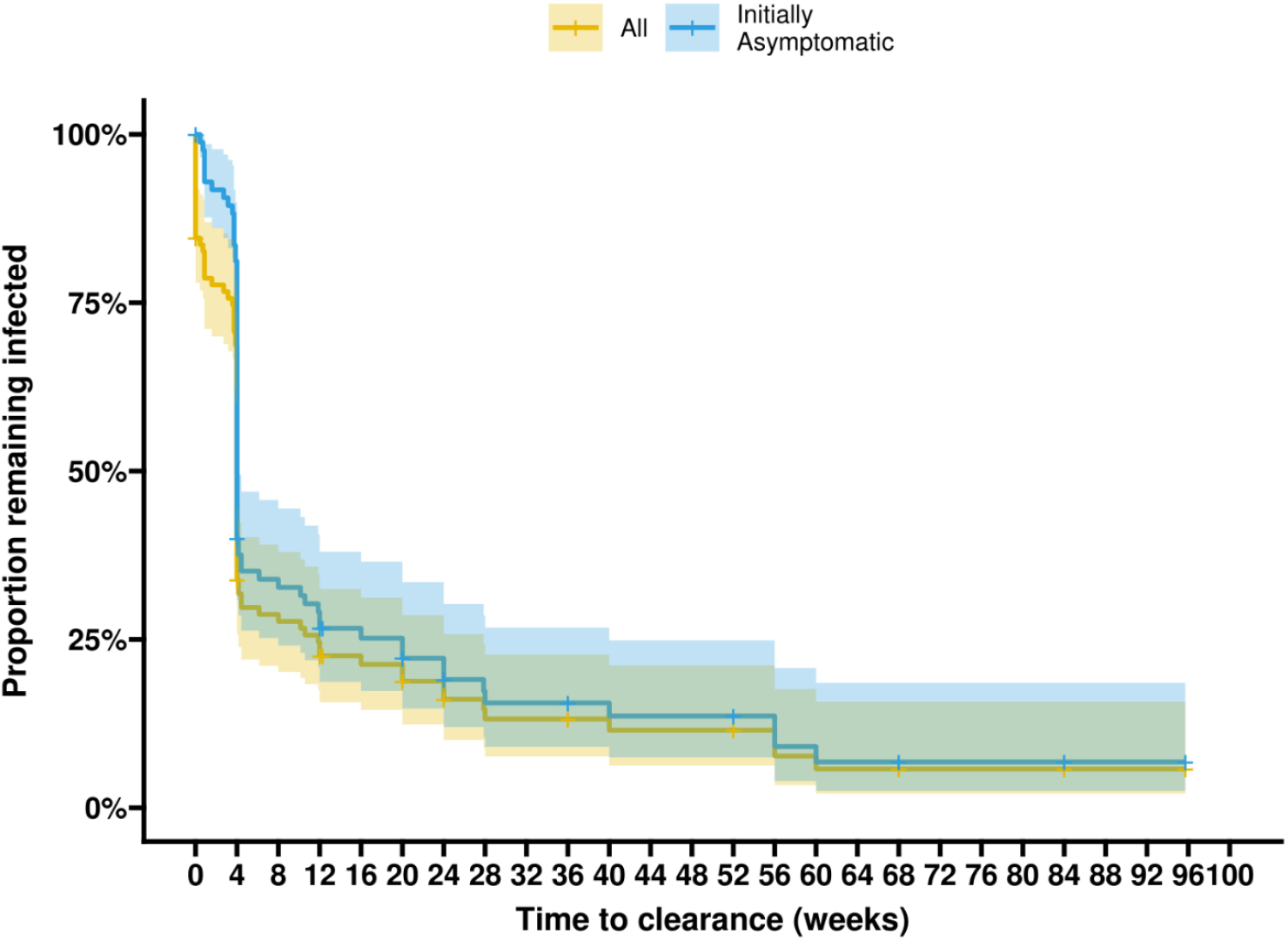
Kaplan-Meier survival curves for all incident infection (yellow) and for initially asymptomatic infections (blue). The y-axis shows the probability of remaining infected (i.e. not clearing parasites) before time in weeks reflected on the x-axis. All 16 initially symptomatic infections were considered cleared, whereas 17.0% (15/88) initially asymptomatic infections (and 14.4% (15/104) of all infections) did not clear by the end of follow-up, with the longest follow-up from the initial detection of infection being 96 weeks. For all infections, we see a steep drop in the proportion at the moment of detection (reflecting the initially symptomatic infections) and again at 4 weeks. 35 infections had cleared at the week visit (all initially asymptomatic and only 2 of which had initiated gametocyte production in the last 4 weeks). After the 4 week visit we see a more conservative but steady decline of the proportion remaining infected (uncleared).

### Most long-duration infections initiate gametocyte production

At first encounter, gametocytes were detected by qRT-PCR in 14.4% (15/104) of all incident infections. Only 6.7% (1/16) of infections that were initially symptomatic and 15.9% (14/88) of infections that were initially asymptomatic had measurable gametocytes at first detection. Competing risks models were used to model time from first detection of an incident malaria infection to the first detection of gametocytes (n=33) while accounting for infection clearance without gametocyte production (n=67) and right-censoring (infections that neither initiated gametocyte production nor cleared throughout follow-up, n=4). In this methodology right-censored observations contribute risk to both event types (gametocyte production or infection clearance without gametocyte production) and thus the estimated proportions may not correspond exactly with observed proportions (which are underestimated because right-censoring is not accounted for). We observed that the likelihood of initiating gametocyte production prior to infection clearance was strongly dependent on the duration of infection. Among initially asymptomatic infections with an infection duration ≥12 weeks, 31.8% (7/22) were observed to have started to produce gametocytes at the first visit (i.e. the infection was detected when first sample detected), while 81.8% (18/22) and 90.9% (20/22) were observed to have initiated gametocyte production 4 and 12 weeks after the infection was detected, respectively. When accounting for right-censoring in these long-duration infections, an estimated 83.3% developed gametocytes by 4 weeks after first detection of infection and 96% developed gametocytes by week 12 (**Figure 2**). Although gametocyte production was thus nearly universal among long-duration infections, these infections were a minority. When considering all initially asymptomatic infections regardless of their duration, only 36.3% (32/88) initiated gametocyte production prior to clearance; 4.5% (4/88) of infections were censored because they neither developed gametocytes nor resolved before study follow-up ended. Accounting for censoring, 29.5% of all infections and 33.7% of all initially asymptomatic infections developed gametocytes within the first month after the infection was detected. Thus, unless gametocytes were produced in the first 28 days, most infections (62.4%) did not produce gametocytes prior to infection clearance (**Figure S2**).

**Figure 2.**
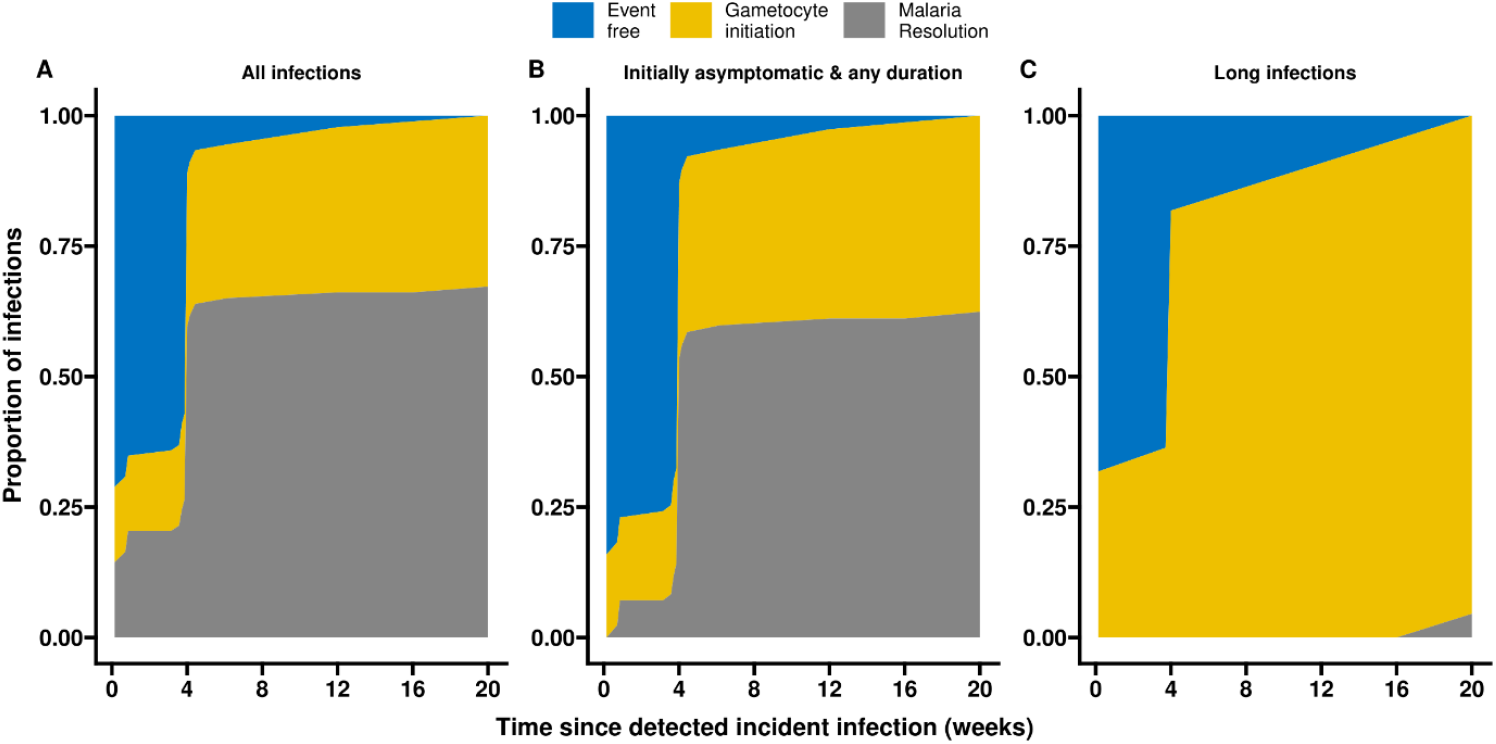
Gametocyte production and infection resolution prior to initiation of gametocyte production over time. These stacked plots show the Aalen-Johansen estimated cumulative proportion of infections that either initiate gametocyte production (yellow) or resolve their malaria without gametocyte production over time -in weeks-(grey). Shown in blue is the proportion of infections that are event free over time, i.e. that have a detectable infection without detectable gametocytes. (A) includes all 104 incident infections, symptomatic and asymptomatic. The percentage of infections initiating gametocyte production at the moment of infection detection (0 weeks) and by 4 weeks was 14% and 29%, respectively. (B) includes 88 infections that were initially asymptomatic. The percentage of infections initiating gametocyte production at 0 and by 4 weeks was 15.9% and 33.7%, respectively. (C) includes 22 infections that were initially asymptomatic and had long duration of infection (≥12 weeks). The percentage of long-duration infections initiating gametocytes by 0, 4 and 12 weeks was 31.8%, 81.8% and 90.9%, respectively.

### Gametocyte density peaks within 4 weeks after infection detection

The geometric mean of peak gametocyte densities was 14.33 gametocytes per µl (IQR 2.56-97.68) among incident asymptomatic infections. Analyzing dynamics of gametocyte densities over the course of infections, 28.1% (9/32) of peak gametocyte densities occurred at the initial visit, 50% (16/32) occurred after the initial visit but by week 4, and 21.9% (7/32) occurred at a timepoint beyond week 4. This temporal pattern of peak gametocyte densities in the first 4 weeks after infection detection, followed by declines in gametocyte densities, was also apparent when average gametocyte densities were plotted per time-interval following infection detection: at the initial visit, 14 initially asymptomatic parasite infections had a geometric mean of 4.61 gametocyte per µl (CI 95%: 0.72-29.51; IQR: 0.58-27.57); this rose to 23.81 (95 CI%: 7.08-80.12; IQR: 13.73-123.47; n=22) gametocyte per µl for observations within the first 4 weeks after infection detection and subsequently declined to a density of 0.39 gametocyte per µl (95% CI: 0.24-0.63; IQR: 0.10-1.35; n=83) across the 12-96 week period post infection detection (**Figure 3**).

**Figure 3.**
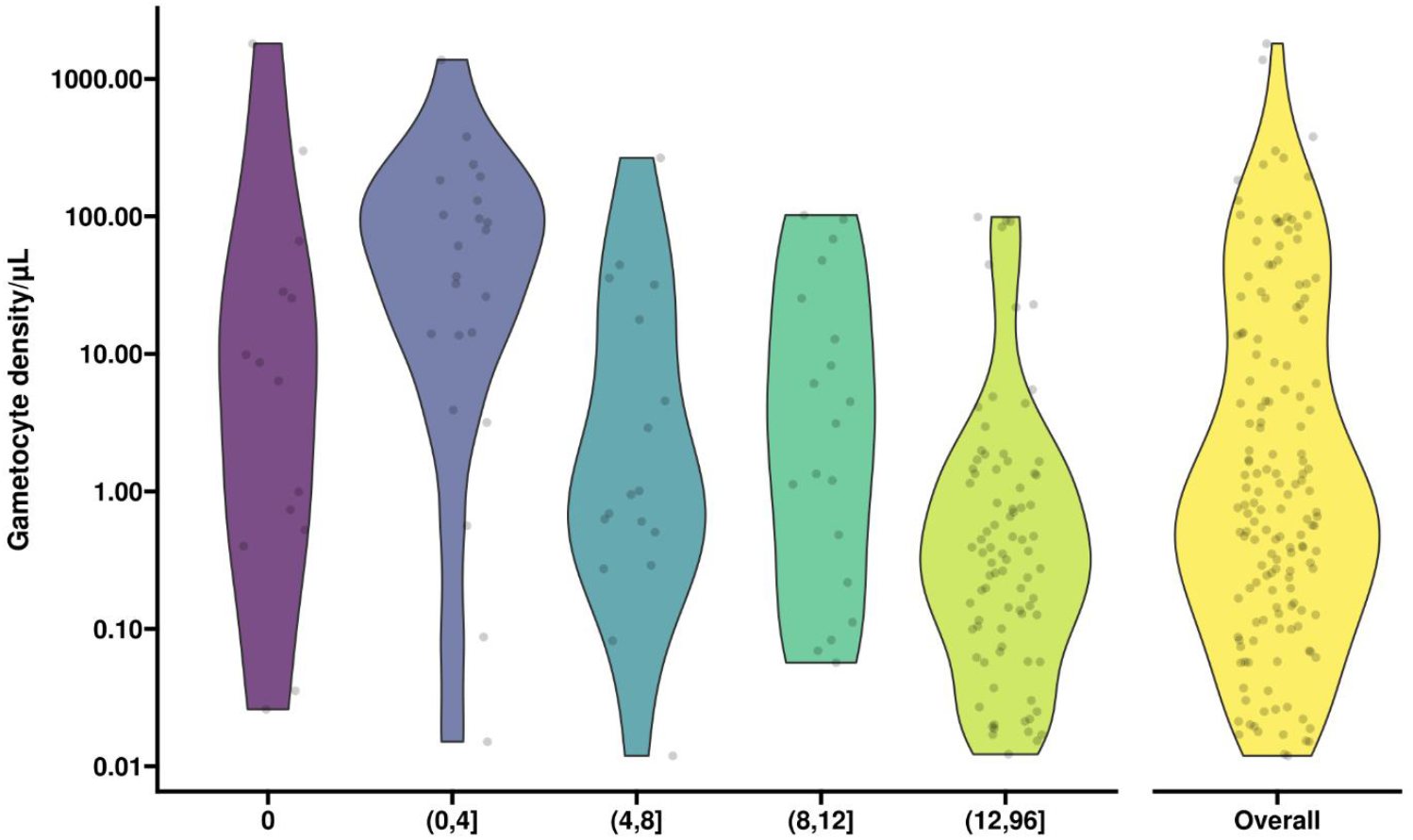
Violin plots of observed gametocyte densities at different time intervals during infections. On the x-axis time in intervals is presented: T=0 moment of infection detection (purple), 0-4 weeks (blue), 4-8 weeks (petrol), 8-12 weeks (green), 12-96 weeks (light green) post infection detection. On the y-axis gametocyte densities per µl are presented. Overall gametocyte density (i.e. over the entire duration of follow-up) is presented in yellow. Gametocyte densities are presented for initially asymptomatic and gametocyte positive observations only (i.e. 32 incident infections with 155 gametocyte positive visits across them). Multiple observations from the same infection may be included in the same violin plot if there were more than one gametocyte positive visit in the interval.

Interestingly, the decline in asexual parasite densities did not mirror that of gametocyte densities. While asexual parasite densities declined immediately following the first detection of infection, followed by a more gradual decline in density after the first 4 weeks of infection, gametocyte densities peaked later (around 4-8 weeks after infection detection) and showed a gradual decline as infections persisted (**Figure 4**). As a consequence of these diverging kinetics, the proportion of parasites that were gametocytes increased over the course of infections (**Figure 4C**).

**Figure 4.**
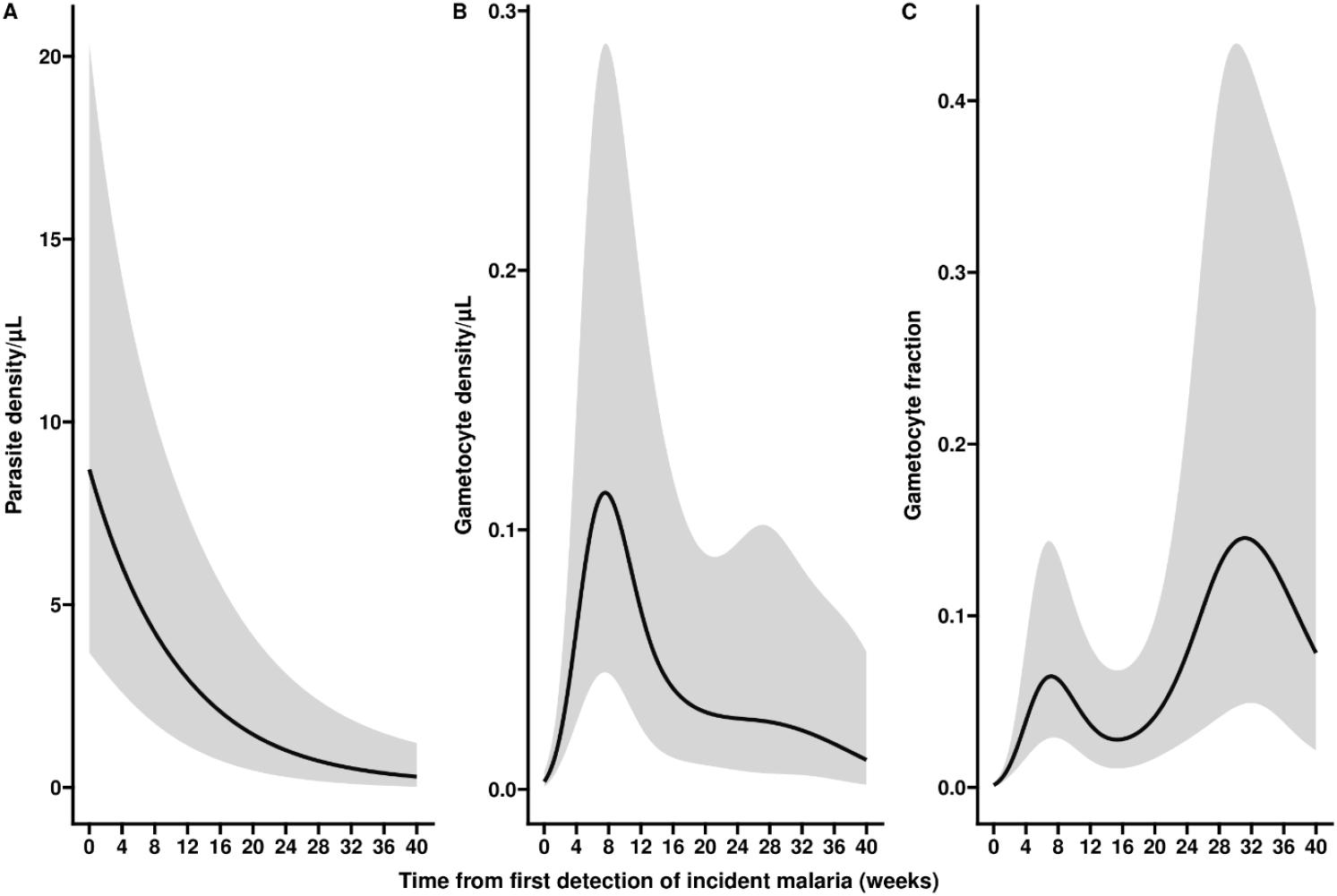
Parasite density, gametocyte density and gametocyte fraction over the course of incident infections. Three separate characteristics of infections are presented: parasite density (**A**), gametocyte density (**B**) and gametocyte fraction (**C**). Gametocyte fraction is defined as the proportion of parasites that are gametocytes, estimated as the proportion of the total parasite biomass (i.e. the density estimated by varATS qPCR) that consists of gametocytes (estimated by Ccp4 and PfMGET qRT-PCR). All estimates are presented over time since first detection of incident malaria; all associations are best described by non-linear mixed-effects models, where a random intercept is used to account for correlation between measurements from the same infection. (**A**) the y-axes shows the estimated parasite density, whilst on x-axis the time from first detection of infection presented in weeks. Parasite density is, on average, at highest levels at the detection of incident malaria and it rapidly declines until ∼ 6 weeks after detection, thereafter a slow and steady decline is observed. (**B**) gametocyte densities are, on average, very low at first detection of incident malaria infections, but rapidly increase until 8 weeks later, thereafter a steady decrease is observed. (**C**) over the course of an infection an increasing trend is seen for the ratio of gametocyte density over parasite density. An estimated decrease is seen at later time-points but this trend is highly uncertain, reflected in wide confidence intervals which also support an increase or a plateau.

### Gametocyte production is dependent on parasite density, multiplicity of infection and sickle cell trait

The initiation of gametocyte production was positively associated with parasite density over the course of infections. Higher parasite densities were associated with higher incidences of gametocyte production (**Figure 5A)**; an apparent plateau in the association between parasite density and the incidence of gametocyte initiation (**Figure 5A**) may be explained by symptomatic infections that are associated with high parasite densities but low incidence of gametocyte initiation due to interruption of the infection by antimalarial treatment.

**Figure 5.**
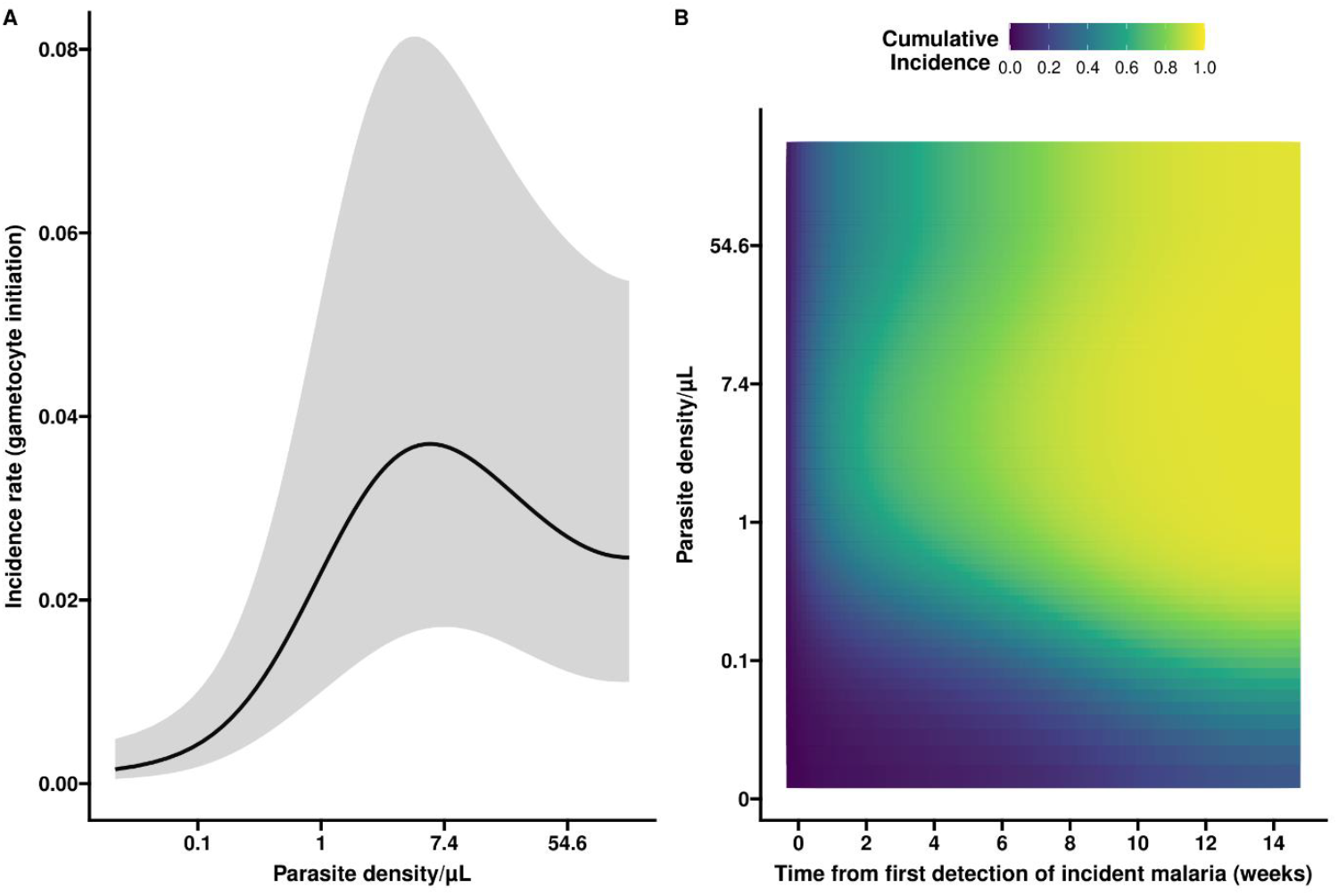
Gametocyte production in relation to parasite density and infection duration. Panel A shows the non-linear association between parasite density/µl (log transformed values) and the incidence of gametocyte initiation (person-day), adjusted for duration of infection. Higher parasite densities are associated with higher incidence rates of gametocyte initiation at a given time until a certain point. While the association is seemingly negative after this point, the wide confidence intervals (grey shaded areas) indicate high levels of uncertainty. Panel B presents the association between parasite density and the duration of incident infection with the cumulative incidence of gametocyte initiation. In Figure 5B, on the y axis the parasite density is expressed per µl, while the x axis describes the time in weeks since the first detection of infection. Low cumulative incidence is shown in blue, whilst cumulative incidences closer to 100% are shown in yellow. Expected cumulative incidences for gametocyte initiation are presented over the full duration of an infection if parasite densities would be maintained at the level given on the y-axis; the likelihood of having initiated gametocyte production increases with increasing parasite density and longer time since infection.

**Figure 5B** illustrates the association between cumulative incidence of gametocyte initiation and parasite density. For this, it is assumed that parasite densities are maintained at constant levels over the course of the infection. If an infection maintains parasite densities above a certain threshold, e.g. at 1 parasites/µL, over time, we expect nearly all incident infections to initiate gametocyte production by 8-10 weeks after detection. The pattern demonstrates that if lower parasite densities are maintained over the course of infection, as was often the case in our cohort, the cumulative incidence of gametocyte production is similarly reduced.

Parasite density was positively associated with gametocyte density but negatively associated with the proportion of parasites that was gametocyte (**Figure 6**). This pattern remained apparent when excluding symptomatic infections (**Supplemental Table 2**). Adjusted for parasite density, initiation of gametocyte production and gametocyte densities over time were associated with several host characteristics. Although not statistically significant, children aged 5-15 years were 1.93 times more likely to initiate gametocyte production over time than children <5 years of age (**Table 2**). Moreover, participants 16 years and older were less likely to resolve their infection before gametocytes were produced, when compared with children under-five (p=0.0001). Notably, individuals with sickle cell trait (HbAS heterozygous individuals) were over two times more likely (Hazard Rate (HR) = 2.68, 95% CI: 1.12, 6.38; p=0.0231) to initiate gametocyte production compared to wildtype (HbAA) individuals over time. In contrast, no association was observed for the limited number of HbSS individuals in our cohort (n=10; HR=1.04, 95% CI: 0.21-5.05; p=0.9637). HbAS individuals also presented with higher gametocyte densities over the duration of infection compared to those with HbAA (Density Ratio (DR) = 9.19, 95% CI: 2.79-30.23; p=0.0002). This elevated gametocyte density for HbAS compared to HbAA individuals greatly exceeded differences between these populations in total parasite density (**Table 2**) and resulted in a 23-fold increase in gametocyte fraction for HbAS parasite carriers, compared to HbAA. The same trend was seen when comparing peak gametocyte densities in HbAS individuals (73.59 gametocytes/µL, IQR: 31.32-189.12) with those in HbAA individuals (4.97 gametocytes/µL, IQR: 0.51-29.29; p=0.0060) and when excluding individuals who were symptomatic at the first moment of infection detection (**Supplemental Table 2**).

**Figure 6.**
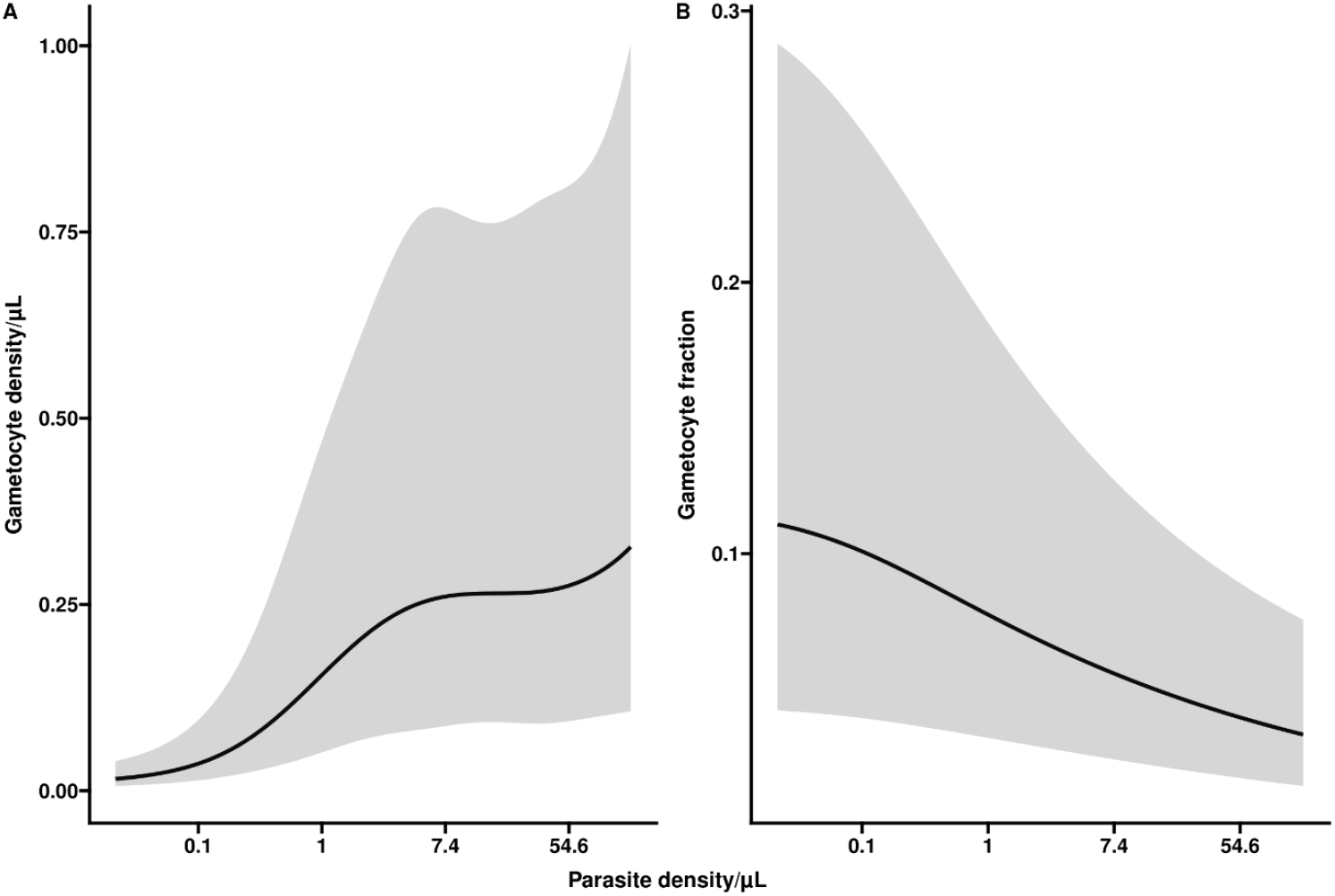
Gametocyte density and gametocyte fraction in relation to parasite density. Parasite density/µl is plotted in log scale within the range of the data. In panel A, the association of parasite density with gametocyte density is presented, adjusted for the duration of infection. We observe a positive association between total parasite density and gametocyte density. In panel B, the association of parasite density with gametocyte fraction is presented, adjusted for the duration of infection. Higher parasite densities are associated with lower gametocyte fraction at any given moment in time. Visits when the infection was detected without detectable gametocytes were included in these analyses.

**Table 2.** Gametocyte production is dependent on parasite density, multiplicity of infection and HbAS. This table describes the effect of host (age, gender, Hb polymorphism) and parasite characteristics (clonality) on the main outcome measures parasite density, incidence of the initiation of gametocyte production (gametocyte incidence), gametocyte density, gametocyte fraction and the incidence of infection clearance without gametocyte production (malaria resolution) with respect to their evolution over time since infection detection. The hazard ratios (HRs) are used to summarize the competing risks modelling of time to two possible events: initiation of gametocyte production or time to parasite clearance without initiating gametocytes. For each event and across time, the hazard rate is defined as the rate at which an infection experiences the event at some time conditional on the infection being event-free before that time. Gametocyte incidence is thus the rate at which infections initiate gametocyte production and malaria resolution is the rate at which infections clear without gametocyte production. The HR for a covariate summarizes how much higher/lower the hazard rate over time is for a covariate value relative to a baseline value for that covariate. The density ratios (DRs) are used to summarize the non-linear mixed-effects modelling of the longitudinal trajectories of parasite density, gametocyte density and gametocyte fraction over time. The DR for a covariate, similar to HR, summarizes how much higher/lower the densities over time is for a covariate value relative to a baseline value for that covariate.

|  | Parasite density |  | Gametocyte incidence |  | Gametocyte density |  | Gametocyte fraction |  | Malaria resolution |  |
| --- | --- | --- | --- | --- | --- | --- | --- | --- | --- | --- |
|  | DR (95% CI) | p-value | HR (95% CI) | p-value | DR (95% CI) | p-value | DR (95% CI) | p-value | HR (95% CI) | p-value |
| **Age (<5 years = reference group)** |  |  |  |  |  |  |  |  |  |  |
| **5-15 years** | 0.72 (0.09, 5.87) | 0.7505 | 1.93 (0.71, 5.23) | 0.1853 | 1.46(0.38, 5.68) | 0.5777 | 1.09 (0.35, 3.38) | 0.8727 | 0.82 (0.43, 1.58) | 0.5471 |
| **16+ years** | 0.12 (0.01, 1.13) | 0.0589 | 1.16 (0.38, 3.54) | 0.7835 | 0.79 (0.19, 3.33) | 0.7447 | 1.12 (0.34, 3.73) | 0.8462 | 0.24 (0.12, 0.49) | 0.0001 |
| **Gender (female = reference group)** |  |  |  |  |  |  |  |  |  |  |
| **Male** | 0.5 (0.09, 2.71) | 0.4133 | 0.97 (0.45, 2.09) | 0.9402 | 1.11 (0.39, 3.17) | 0.845 | 1.24 (0.53, 2.89) | 0.6165 | 0.75 (0.43, 1.31) | 0.3055 |
| **HB genotype (HbAA = reference group)** |  |  |  |  |  |  |  |  |  |  |
| **HbSS** | 8.33 (0.45, 154.16) | 0.1464 | 1.04 (0.21, 5.05) | 0.9637 | 1.53 (0.23, 10.11) | 0.6512 | 1.55 (0.31, 7.88) | 0.5869 | 1.44 (0.61, 3.39) | 0.3952 |
| **HbAS** | 1.69 (0.22, 13.11) | 0.6083 | 2.68 (1.12, 6.38) | 0.0231 | 9.19 (2.79, 30.23) | 0.0002 | 3.13 (1.14, 8.59) | 0.0241 | 0.43 (0.19, 0.96) | 0.036 |
| **Clonality (monoclonal infection = reference)** |  |  |  |  |  |  |  |  |  |  |
| **Multiclonal** | 15.43 (2.62, 90.98) | 0.002 | 2.93 (1.16, 7.44) | 0.0208 | 5.96 (1.4, 25.32) | 0.0135 | 2.45 (0.76, 7.87) | 0.1255 | 0.40 (0.17, 0.91) | 0.0265 |

Regarding infection characteristics, polyclonal infections were associated with an increase in both asexual parasite densities (15-fold) and in gametocyte densities (6-fold) (**Table 2**). As a result, there was no measurable increase in gametocyte fraction in polyclonal infections (p=0.1255). Further supporting the role of HbAS and polyclonal infections in gametocyte production, we observed that HbAS individuals and individuals with polyclonal infections were less likely to resolve their infection prior to initiating gametocyte production (p=0.0360 and p=0.0265, respectively); these associations remained significant after adjusting for parasite density at first detection of infection. The limited number of incident infections with information on the complexity of infection, made it impossible to explore the effects of sickle cell status and multiplicity of infection concurrently in a multivariate model.

### The associations were not driven by incident infections with very low parasite densities

We performed a sensitivity analysis by removing all 29.8% (31/104) of infections with an initial parasite density below 0.1 parasites per µl and re-running the models (**Supplemental Table 1**). The evolution of parasite density, gametocyte density and gametocyte fraction remained the same but, as a consequence there was a shift in the positive direction on the y-axis for parasite density and gametocyte density (not for gametocyte fraction) (**Supplemental S5**), driven by the higher parasite densities. The association between parasite density and gametocyte initiation incidence showed a similar trend to **Figure 5**, with a more apparent plateau (**Supplemental S7**). Similarly, the positive association between parasite density and gametocyte density and the negative association between parasite density and gametocyte fraction remained apparent after excluding infections with low initial parasite densities (**Supplemental S6**). The effects of HbAS and polyclonal infections were similarly consistent (**Supplemental Table 1**).

### The association of polyclonality with gametocyte density may be driven by symptomatic infections

A further sensitivity analysis was performed. Here we removed all 27% (28/104) of infections that were symptomatic at any time. The evolution of parasite density, gametocyte density and gametocyte fraction continued to be the same but, as expected, parasite and gametocyte densities were much lower (**Supplemental S9**). The association between parasite density and gametocyte initiation incidence showed a similar trend to **Figure 5**, (**Supplemental S11**), albeit with much lower cumulative incidences (driven by the lower parasite and gametocyte densities). The positive association between parasite density and gametocyte density and the negative association between parasite density and gametocyte fraction remained persistent (**Supplementary S10**). HbAS was still statistically significantly associated with gametocyte production initiation and gametocyte density, while the effects of having a polyclonal infection on gametocyte density no longer achieved statistical significance (**Supplemental Table 2**).

In addition to the findings of the above sensitivity analysis, we found through a further sensitivity analysis that when accounting for interval-censoring, the effect of polyclonality on time to gametocyte initiation was no longer statistically significant. This interval censoring was considered important since the exact time at which the incident malaria infection, initiation of gametocyte production, or malaria clearance occurred was not known due to the nature of routine sampling every 4 weeks. Ignoring interval-censoring may lead to underestimated standard errors and thus inflated type I error rates, i.e. false positive findings. We investigated how sensitive the results of our modelling were to the interval-censored nature of the event times by imputing exact times using a uniform distribution from the intervals in which they were observed. We imputed one hundred datasets and re-ran the models for all outcomes. For the time to initiation of gametocytes, HbAS showed statistical significance (p<0.05) in 81% of the imputed data, and the remaining 19% were significant at a 10% threshold for significance (p<0.10) (**Supplementary S13 B plot**). Conversely, polyclonality was only statistically significant (p<0.05) in 10% of the imputed data, with a wider range of p-values. For the longitudinal modelling of parasite density, gametocyte density and gametocyte fraction, we see a nearly unanimous agreement across imputations indicating a statistically significant effect of HbAS on gametocyte density and gametocyte fraction and a significant effect of polyclonality on parasite density and gametocyte density. None of the imputations showed a significant effect of HbAS on parasite density or of multiplicity of infection on gametocyte fraction, as we had expected from our main findings.

## Discussion

Here, we longitudinally monitored 104 infections acquired in an area with a low force of infection but historically exposed to intense malaria transmission. We observed that incident infections, that occurred in all age groups, were typically of very short duration and not associated with the production of gametocytes. In the minority of infections that persisted for 3 months or longer, gametocyte formation was near universal with earlier and higher gametocyte production in higher density infections and individuals with sickle cell trait (HbAS).

Infections with *P. falciparum* can be of very long duration. In our cohort, some individuals carried infections for 2 years or longer (Briggs et al., 2020; Andolina et al., 2021) and infections of up to 14 years have been documented elsewhere (Ashley et al., 2014). Because chronic infections are more likely to be detected in community surveys and are considered important sources of transmission (Barry et al., 2021) and plausible hurdles for malaria elimination initiatives (Ashley et al., 2014), they understandably received considerable attention. Our detailed parasitological monitoring with sensitive molecular diagnostics, however, uncovered that only a minority of incident bloodstream infections result in chronic parasite carriage. There are several reports of the occurrence of brief episodes of parasitemia; these are typically based on microscopy and leave uncertainties about possible persistence of parasitemia at submicroscopic densities (reviewed in (Roe et al., 2022)). Using a highly sensitive qPCR (Hofmann et al., 2018) in cohort participants who were initially free of infection, we observed that 62.5% of all incident infections were observed only once. The exact duration of these infections was impossible to estimate with precision, given the 4-week interval between scheduled visits, but it is evident that the majority of infections were brief and spontaneously cleared. While naturally acquired immunity has been suggested as determinant of infection clearance (Bruce et al., 2000; Felger et al., 2012; Nguyen et al., 2018), recent analyses suggest that infection duration may be shortest in young children (Briggs et al., 2020) and acquired immunity may actually be a prerequisite to sustain chronic infections (Collins et al., 2022), rendering a role of immunity-dependent infection clearance less obvious. Importantly, we provide the first evidence that these short-duration infections do not result in measurable gametocyte densities. It is possible that very brief episodes of gametocytemia were missed but we consider it unlikely that this was common, given that gametocytes typically persist for approximately one month after infections are cleared with gametocyte-permissive drugs in controlled human infections (Collins et al., 2018; Reuling et al., 2018). It is, however, conceivable that ultra-low gametocyte densities have been missed. The majority of infections that were of short duration were of very low density and since gametocyte densities comprise only a small proportion of the total parasite biomass, very low gametocyte densities may have remained undetected. The strong association we report between asexual parasite density and initiation of gametocyte production further demonstrates that the detectability of gametocytes is closely linked to that of their asexual progenitors. Our findings are therefore not contradicting the notion of stochastic gametocyte commitment in a minority of parasites (Llora-Batlle et al., 2020) but rather demonstrate that gametocyte densities that can plausibly result in onward transmission to mosquitoes, above ∼5 gametocytes/µL (Bradley et al., 2018; Andolina et al., 2021), are not achieved by the majority of incident infections.

Intriguingly, we observed strong indications for increased gametocyte production in individuals with sickle cell trait (HbAS). HbAS has previously been associated with lower parasite densities and a reduced chance of clinical disease upon infection (Gong et al., 2012; Lopera-Mesa et al., 2015); also an increased transmission potential of HbAS parasite carriers has been suggested before (Gouagna et al., 2010; Lawaly et al., 2010). Our study is the first to quantify enhanced gametocyte formation in relation to HbAS in longitudinally monitored infections. We observed that the initiation of gametocyte production was faster and gametocyte densities over the course of infections were higher for HbAS individuals. Moreover, the proportion of parasites that were gametocytes was higher, suggestive of higher per-parasite gametocyte production. More frequent sampling is required to formally demonstrate differences in gametocyte production but it is conceivable that gametocyte formation is triggered in HbAS blood cells where parasite growth is reduced (Pasvol et al., 1978) and erythrocyte sickling upon parasitization is increased (Luzzatto et al., 1970). Our previously reported assessment of gametocyte infectivity (Andolina et al., 2021) allowed us to explore the viability of gametocytes in HbAA and HbAS carriers. Although data were too sparse for detailed analysis, the representation of different HbS genotypes among infectious individuals (n=21) was similar to that of the entire population (**Table 1**). Among infectious individuals, 23.8% (5/21) had the HbAS genotype and 9.5% (2/21) had HbSS and there was no obvious difference in the association between gametocyte density and mosquito infection rate (**Figure S4**). Whilst we found no evidence to support previous claims of higher gametocyte infectivity in HbAS carriers (Trager et al., 1999; Gouagna et al., 2010), our mosquito feeding assays give us confidence that our observation of higher gametocyte production translates in an increased transmission potential for HbAS carriers. This increased transmission potential will be aggravated by the fact that infections also last longer in HbAS carriers (Gong et al., 2012).

The presence of competing parasite clones is also considered as trigger for increased gametocyte formation (Nassir et al., 2005; Lamptey et al., 2018; Sondo et al., 2021). Our findings of higher gametocyte densities in polyclonal infections appear supportive of these associations but nevertheless have to be interpreted with caution. Although patterns persisted in a sensitivity analysis where the lowest-density infections were excluded, it remains possible that the association is (partly) driven by a higher detectability of parasite clones in higher density infections that are also associated with higher gametocyte production.

In this study, we modelled the non-linear trajectories of parasite density, gametocyte density, gametocyte incidence and gametocyte fraction over time using the generalized additive mixed modelling (GAMM) framework (Wood et al., 2016; Wood, 2017). To model the effects of covariates, we were able to adjust for this non-linear effect of parasite density over time. GAMMs allows for the modelling of complex non-linear relationships using smoothing splines (Belias et al., 2022) without the need for parametric assumptions of the shapes of the trajectories. GAMMs are extensively used in areas such as ecology (Pedersen et al., 2019), and have been used to model other infectious disease processes with complex non-linear effects and interactions, such as seasonality (Ramjith et al., 2021). While under-utilized in malaria epidemiology, Rodriguez-Barraquer (Rodriguez-Barraquer et al., 2018) used GAMMs to model the malaria incidence, parasite prevalence and density over age and exposure. They showed how the non-linear trajectories of malaria incidence and parasite prevalence over age varied across sites and they showed the complex bivariate effect of age and exposure on parasite density and the ‘fever threshold’.

One relevant limitation of our work is that the exact time of incident infection, and gametocyte initiation was not known. The generalized additive mixed modelling (GAMM) framework (Wood et al., 2016; Wood, 2017) that we used to model the non-linear trajectories of parasite density, gametocyte density, gametocyte incidence and gametocyte fraction over time, does not account for the uncertainty surrounding observation of the exact times. Here, duration of infection was defined as the time from first detected incident infection and for the time-to-event models, the event time was defined as the time from the first detected incident infection to first detected gametocyte production. To investigate how sensitive our model was to the interval censored nature of the incident infection, and also for the start of gametocyte production and infection resolution (for the time-to-event model), we imputed 100 datasets with times uniformly generated from within their intervals and observed consistency in our reported associations between gametocyte metrics and HbAS and polyclonality of infections. GAMMs allow for the modelling of complex non-linear relationships using smoothing splines (Belias et al., 2022) without the need for parametric assumptions of the shapes of the trajectories. GAMMs are extensively used in areas such as ecology (Pedersen et al., 2019), and have been used to model other infectious disease processes with complex non-linear effects and interactions, such as seasonality (Ramjith et al., 2021). As a result of the limited number of infections and failure to determine the complexity of infection in a substantial number of infections, especially in low-density infections (Early et al., 2019), we were unable to assess the impact of covariates on gametocyte production in a multivariate model.

In conclusion, this study provides evidence that a large proportion of infections in all age groups are of short duration and low parasite density, lasting less than a month, and not associated with gametocytemia. Although in minority, long lasting chronic infections are characterized by high gametocyte production and transmission potential and infection duration and gametocyte production were influenced by host genetic status. These findings, from an area where intensive mosquito control which was exceptionally effective in driving down transmission, are of relevance for malaria elimination initiatives. Such initiatives have to consider that whilst chronic asymptomatic infections form a major reservoir for infectious gametocytes, this status is only achieved by a minority of incident infections.

## Material and methods

### Study site

Data were obtained from a prospective cohort study conducted in Nagongera subcounty, Tororo district near the Kenyan border between October 2017 and October 2019 (Nankabirwa et al., 2020). Historically, this was an intense transmission setting where malaria transmission was highly reduced following repeated long-lasting insecticidal nets (LLIN) distributions and sustained and effective and indoor residual spraying of insecticides (IRS) (Nankabirwa et al., 2020). 531 participants were recruited from 80 randomly selected households as described previously (Andolina et al., 2021). At enrollment, blood samples were screened for sickle cell trait HbAS.

Cohort study participants were encouraged to come to a dedicated study clinic open 7 days per week for all their medical care. Routine visits were conducted every 28 days and included a standardized clinical evaluation and collection of blood for thick blood smear, hemoglobin measurement (every 12 weeks), and storage for future molecular studies including ultrasensitive varATS qPCR and gametocyte quantification by reverse transcriptase qPCR targeting female (Ccp4) and male (PfMGET) gametocyte transcripts (Kamya et al., 2015). Study participants with fever (tympanic temperature > 38.0°C) or history of fever in the previous 24 hours had a thick blood smear read immediately. If the thick blood smear was positive by light microscopy, the patient was diagnosed with malaria and managed according to national guidelines (Uganda, 2016). Participants with asymptomatic parasitemia were not given antimalarial treatment in accordance with national guidelines.

### Laboratory methods

200 µL whole blood samples were collected at enrollment, at routine each visit and at each clinical malaria episode (Nankabirwa et al., 2020). DNA extraction was performed by Qiagen spin columns (Qiagen QIAamp DNA Blood Mini Kit) and 5 µl of extraction products was assessed for presence and quantification of *P*.*falciparum* parasites by varATS qPCR which has a limit of detection of 0.05 parasites/µL (Hofmann et al., 2015). Samples with >0.1 parasite/µl were genotyped by apical membrane antigen-1 (AMA-1) amplicon deep sequencing (Briggs et al., 2020).

For all qPCR positive samples, 100 µL of blood in RNA protect (RNA protect Qiagen), prior to DNA extraction, was used to quantify female (CCp4 mRNA) and male (PfMGET mRNA) gametocyte transcripts by quantitative reverse transcriptase PCR (qRT-PCR) with a lower limit of detection of 0.1 gametocytes/µL (Meerstein-Kessel et al., 2018).

### HBS genotyping

Genotyping of HbS mutation (dbSNP rs334) used molecular inversion probe (MIP) capture and deep sequencing on Illumina NextSeq 550 (Aydemir et al., 2019). MIP capture incorporated unique molecular identifiers (UMIs) allowing subsequent collapse of reads representing PCR duplicates of the same captured molecule. Individuals SNP genotype calls required >=10 UMI depth and >=Q20 genotype quality to ensure capturing both alleles (99.8%) in a heterozygous individual with trait.

### Gametocyte outcome measures

We analyzed main outcome measures such as the incidence of the initiation of gametocyte production and the incidence of infection clearance without gametocyte production, parasite density, gametocyte density and gametocyte fraction with respect to their evolution over time since infection detection and with respect to host and parasite characteristics. The incidence of the initiation of gametocyte production describes the measure of the frequency of first detection of gametocytes over time whilst the incidence of the infection clearance prior to gametocyte production describes the frequency of infections that resolve without gametocyte production. The incidence of the initiation of gametocyte production or infection clearance without gametocytes were measured through the hazard rate, defined as the likelihood of an event occurring over time conditional that it did not occur before. In addition we also estimated the cumulative incidences, i.e. the proportion of infections that initiated gametocytes before a certain time and the proportion of infections that cleared without gametocyte production by a certain time. This is different from the observed proportions, in that the observed proportions exclude right-censored observations entirely after the time in which they are right-censored. These right-censored observations were incident infections that did not initiate gametocyte production and did not clear by the end of their follow-up. The gametocyte fraction was defined as the proportion of the total parasite biomass of asexual ring stages which was gametocyte.

### Incidence model: detection of gametocyte initiation over time since first detection of infection

The time from first detection of an incident infection with malaria parasites to the detection of gametocytes was studied applying theory from survival analysis. In survival analysis, this time-span is often modeled via the hazard function which is a function of time as well as covariates of interest (e.g. parasite characteristics, participant demographic or genetic characteristics). Here, we chose to model this hazard function as a piece-wise exponential additive model (PAM) to have sufficient flexibility (Argyropoulos et al., 2015; Bender Andreas, 2018). In the analysis, a competing risk needed to be accounted for since some incident infections may resolve before gametocyte production is initiated and these incident infections no longer contribute risk for initiating gametocyte production. Therefore, we considered the *cause-specific* baseline hazards functions for time to gametocyte initiation and for time to clearance of infection prior to gametocyte initiation. The parameters in the model were estimated by a penalized maximum likelihood method within a competing risks framework. More specifically, and also similarly described elsewhere (Kopper Philipp; Ramjith, 2021; Ramjith et al., 2021; Bender Andreas, 2018) the hazard function at time *t* is defined as

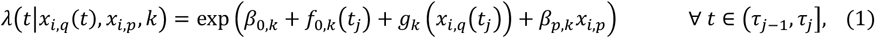

where events *k* = 1, 2 indicate gametocyte initiation and clearance of infection prior to gametocyte initiation respectively, exp (*β*_*0,k*_ + *f*_*0,k*_(*t*_*j*_)) is the baseline hazard function at time *t*_*j*_ for each of the events respectively, and *t*_*j*_ is a chosen value of time within the interval (in our case τ _*j*_ = *τ*_*j*_, the right boundary) that was used to make the hazard depend on time while being constant over time within the interval. Note that other choices of *t*_*j*_ within the interval (*τ*_*j−*1_, *τ*_*j*_) can be made. Furthermore, *g*_*k*_(.) is the non-linear function for the effect of the time-varying covariate log parasite density (*x*_*i,q*_(*t*)) for each of the *k* events. Regression coefficient β_*p,k*_ is the unknown coefficient for the effect of the *p*^*th*^ covariate *x*_*p*_ for each of the *p* covariates. Non-linear functions are represented through basis functions and coefficients, e.g. 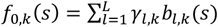 where*γ*_*l,k*_ are unknown regression parameters to be estimated and *b*_*l,k*_(*s*) are chosen basis functions. We used B-spline basis functions (Deboor, 1978). A succinct introduction to splines, including B-splines, is provided by (Belias et al., 2022). The PAM applied to the time intervals is equivalent to the Poisson GAMM where the effect of time is modelled with penalized splines stratified by event type, but without the random-intercepts component. This makes it possible to estimate the parameters in the model by their maximum likelihood estimates. To overcome overfitting a penalty was added to the log likelihood function.

We considered several models. We first fit a model with only a spline for time to visualize the baseline hazards for both events, gametocyte initiation and infection clearance prior to gametocyte initiation, i.e. λ(*t*|*k*) = exp (*β*_*0,k*_ + *f*_*0,k*_(*t*_*j*_)). To visualize the (potentially non-linear) effect of parasite density as a time-varying covariate on the incidence of gametocyte initiation, we fit a model with a penalized spline for the non-linear effect of log parasite density, i.e. *λ*(*t*|*x*_*i,q*_(*t*), *k*) = exp (*β*_*0,k*_ + *f*_*0,k*_(*t*_*j*_) + *g*_*k*_ (*x*_*i,q*_(*t*_*j*_))). To assess the effects of the remaining time independent categorical covariates, we fit independent models for each covariate as specified in (1).

### Non-linear models: Parasite density, gametocyte density and gametocyte fraction over time since first detection of infections

To study the non-linear trajectories of the repeated measurements of parasite density, gametocyte density and gametocyte proportion (proportion of the total parasite biomass that is gametocyte) over the duration of infection we fit generalized additive mixed models (GAMMs) for each of these log transformed outcomes respectively.

Random intercepts were included in the model to account for the correlation between observations within the longitudinal profile of the infection for an individual. More specific, for individual *i* the (conditional) expected logarithm of the outcome (denoted as *E*(*log y*_*ijk*_ |*z*_*i*_), where *k* = 1,2,3 for parasite density, gametocyte density and gametocyte fraction respectively) at time point *t*_*j*_ is assumed to be equal to:

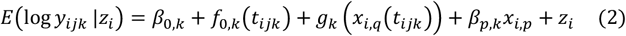

where *k* = 1, 2, 3 define the outcomes parasite density, gametocyte density and gametocyte fraction respectively, *f*_*0,k*_(.) is the non-linear function which expresses the effect of infection duration and *g*_*k*_(.) is the non-linear function for the effect of the time-varying covariate log parasite density (*x*_*i,q*_(*t*)) for outcomes gametocyte density and gametocyte fraction (*k* = 2, 3) and 0 otherwise (i.e. *g*_1_(.) = 0). Regression coefficient *β*_*p,k*_ is the unknown coefficient for the effect of the *p*^*th*^ covariate *x*_*i,p*_ for each of the *p* covariates (for individual *i*) and *z*_*i*_ is the random intercept for the *i*^*th*^ individual. Non-linear functions are represented through basis functions and coefficients, e.g. 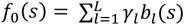 where *γ*_*l*_ are unknown regression parameters to be estimated and *b*_*l*_(*s*) are chosen basis functions. B-spline basis functions were used.

We first fit models with only the penalized spline for time for each outcome to visualize the baseline expected parasite density, gametocyte density and gametocyte fraction over the duration of infection, i.e. *E*(*log y*_*ijk*_ |*z*_*i*_) = *β*_*0,k*_ + *f*_*0,k*_(*t*_*ijk*_) + *z*_*i*_. To visualize the (potentially non-linear) effect of parasite density as a time-varying covariate on gametocyte density and gametocyte fraction, we fit a model with a penalized spline for the non-linear effect of log parasite density, i.e. *E*(*log y*_*ijk*_ |*z*_*i*_) = *β*_*0,k*_ + *f*_*0,k*_(*t*_*ijk*_) + *g*_*k*_ (*x*_*i,q*_(*t*_*ijk*_)) + *z*_*i*_, for *k* = 2, 3. To assess the effects of the remaining time invariant categorical covariates on each for the outcomes, we fit independent models for each covariate as specified in (2).

We present the results for the time-invariant categorical covariates as density ratios (DRs): *DR* = exp *β*_*p,k*_ which quantifies how much larger (multiplicatively) the outcome (parasite and gametocyte densities or gametocyte fraction) is for all levels of the *p*^*th*^ covariate *x*_*p*_ relative to its reference category.

## Data Availability

Data from the PRISM2 cohort study is available through a novel open-access clinical epidemiology database resource

https://clinepidb.org/ce/app/record/dataset/DS_51b40fe2e2.

## Data and software availability

Data from the PRISM2 cohort study is available through a novel open-access clinical epidemiology database resource here: https://clinepidb.org/ce/app/record/dataset/DS_51b40fe2e2.

## Acknowledgments

We thank all the study participants for their willingness to support the study, attend the monthly visits and donate blood. We also thank all the lab staff, drivers and procurement for their dedication in the study.

## Additional information Funding

Funding was provided by the National Institutes of Health as part of the International Centers of Excellence in Malaria Research (ICEMR) program (AI089674) and another program (AI075045), the Bill & Melinda Gates Foundation (INDIE OPP1173572) and a fellowship from the European Research Council to TB (ERC-CoG 864180; QUANTUM). BG is a Chan Zuckerberg Biohub investigator. The ClinEpiDB platform is supported by the Bill & Melinda Gates Foundation (OPP1169785).

## Author contributions

Chiara Andolina, Conceptualization, Writing-original draft, Data curation; Jordache Ramjith, Conceptualization, Formal analysis, Visualization, Data curation, Writing-original draft, Methodology; John Rek, Supervision, Project administration; Kjerstin Lanke, Methodology; Joseph Okoth, Supervision; Lynn Grignard, Methodology; Emmanuel Arinaitwe, Supervision, Project administration; Jessica Briggs, Methodology; Jeffrey Bailey, Methodology, Writing - review and editing; Ozkan Aydemir, Methodology; Moses R Kamya, Resources, Supervision, Funding acquisition, Project administration, Writing - review and editing; Bryan Greenhouse, Resources, Funding acquisition, Supervision, Writing - review and editing; Grant Dorsey, Resources, Funding acquisition, Supervision, Writing - review and editing; Sarah G Staedke, Resources, Supervision, Writing - review and editing; Chris Drakeley, Supervision, Funding acquisition, Writing - review and editing; Marianne Jonker, Conceptualization, Supervision, Writing-original draft, Writing - review and editing; Teun Bousema, Conceptualization, Resources, Supervision, Funding acquisition, Writing-original draft, Writing - review and editing

