## Supplementary material for "Gametocyte production in incident *P. falciparum* infections: a longitudinal study in a low transmission setting under intensive vector control"

**Supplemental figures**

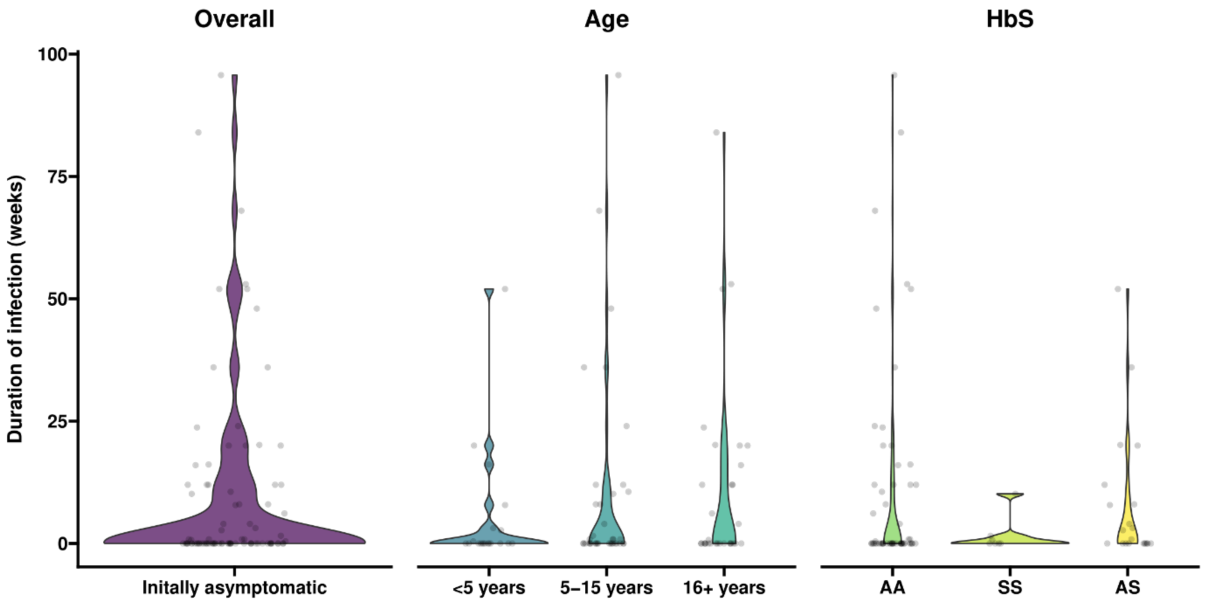


**Figure S1**: **Violin plots of duration of infection in initially asymptomatic (purple), in different age groups <5, 5-15, 16+ years old (blue, petrol, turquoise) and in Hb AA, SS, AS (green, light green, yellow).** There was no association between duration of infection with age (p=0.5049) and Hb (p=0.6648).

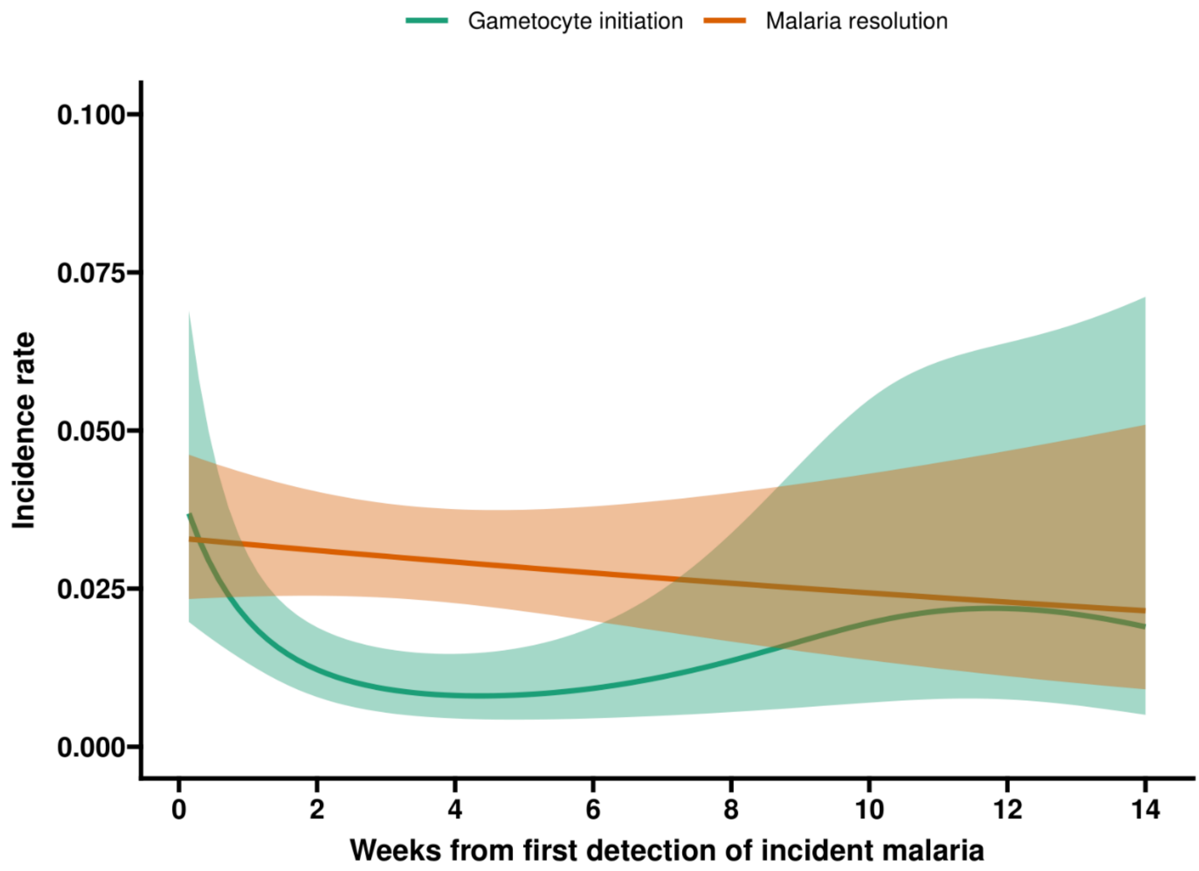


**Figure S2: Baseline incidence density/hazard rates for gametocyte production and resolution without initiating gametocytes over the time since detected incident malaria infection.** This figure illustrates the unadjusted baseline plots of how the incidence of gametocyte initiation and also malaria resolution prior to gametocyte initiation evolves over time since detected infection. The incidence rate is the probability of experiencing either of the events over time since detection among those who are still event free at that time. The largest incidence of gametocyte production occurred soon after detection of incident malaria infections (green line) and decreased overtime. Whilst the incidence rate of malaria resolution steadily decreased over the duration of infection.

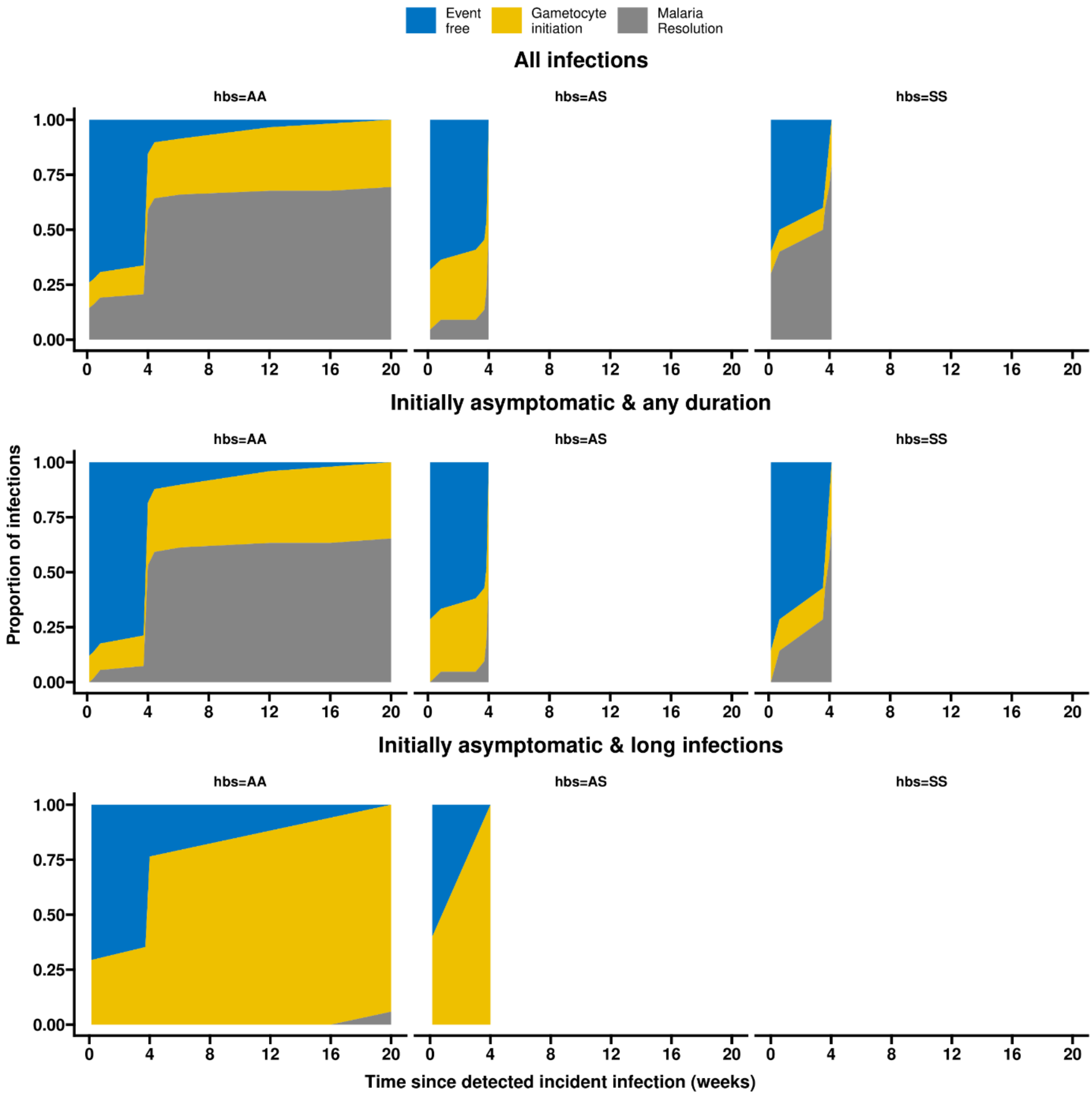


**Figure S3: Gametocyte production and infection resolution prior to initiation of gametocyte production over time.** This stacked plot shows the cumulative proportion of infections that either initiate gametocyte production (yellow) or resolve their malaria without gametocyte production over time (grey). Shown in blue is the proportion event free over time. All plots are stratified for HbS where AA (wild type) is shown on the left panel, AS (heterozygote) in the middle panel and SS (homozygote) on the right panel. The top panel is for all 104 infections, of which 3 had missing HbS data, 69 were AA, 10 SS and 22 AS.  By 4 weeks approximately half the AS infections initiated gametocytes and the other half resolved. The middle panel is for the 24 infections that were initially asymptomatic and had >12 weeks total duration of infection, of which 17 were AA, 1 SS and 6 AS. 76% (13/17) AA initiated gametocytes by 4 weeks and 100% (6/6) AS initiated gametocytes by 4 weeks. The bottom panel is for the 88 infections that were asymptomatic initially and for all durations of infection, of which 2 HbS data were missing, 58 were AA, 7 SS and 21 AS. 26% (15/57) AA and 52% (11/21) AS infections initiated gametocytes by 4 weeks.

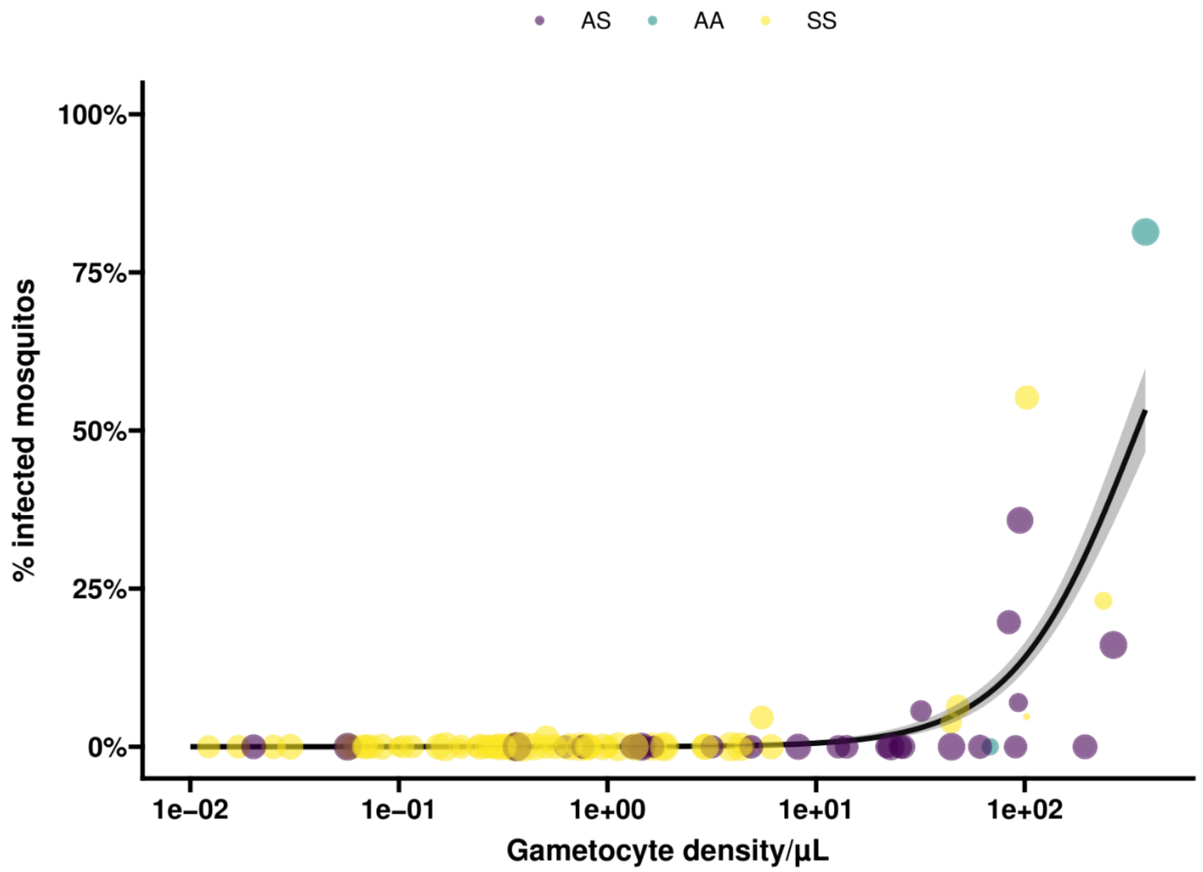


**Figure S4**: **Percentage of infected mosquitoes in relation to gametocyte density expressed per µl for AS (purple), AA (petrol), SS (yellow) individuals**. The size of the dots reflects the number of mosquitoes dissected.

**Sensitivity analysis-parasite density cut off <0.1 parasites per µl**

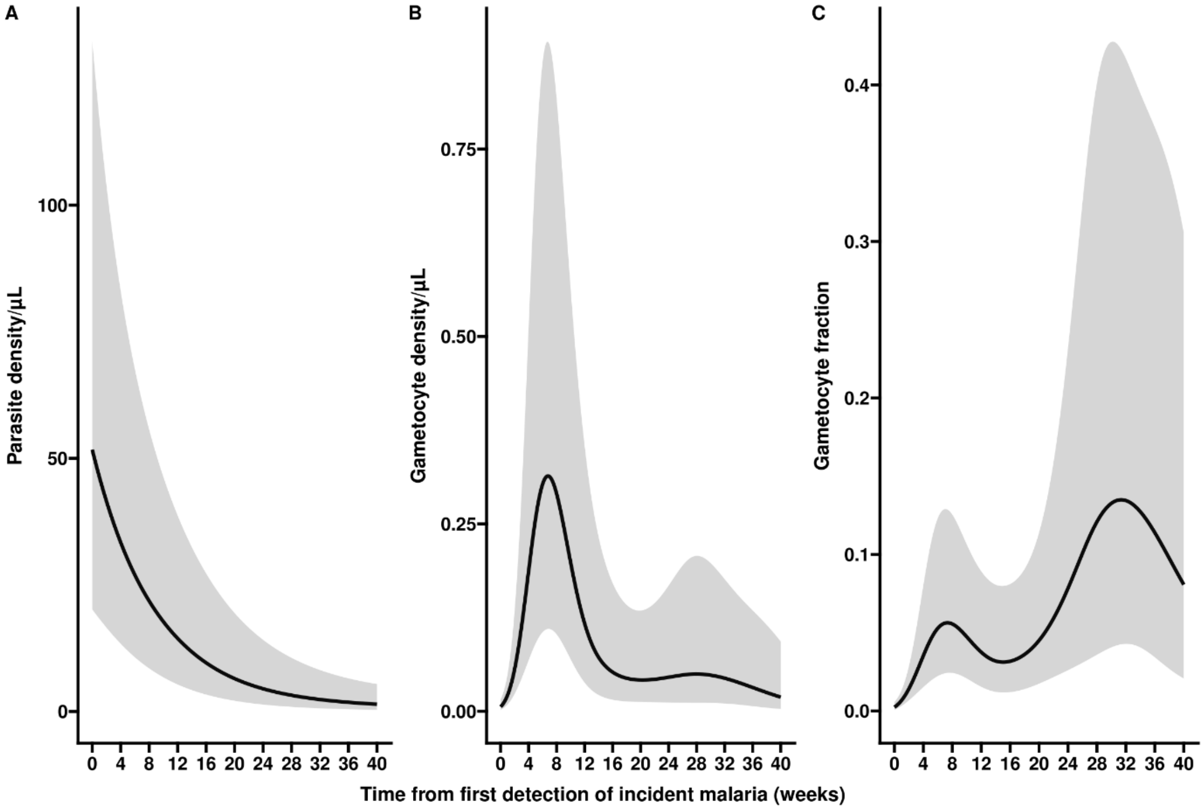


**Figure S5 Parasite density, gametocyte density and gametocyte fraction over the course of incident infections.** Three separate characteristics of infections are presented: parasite density (A), gametocyte density (B) and gametocyte fraction (C). Gametocyte fraction is defined as the proportion of parasites that are gametocytes, estimated as the proportion of the total parasite biomass (i.e. the density estimated by varATS qPCR) that consists of gametocytes (estimated by Ccp4 and PfMGET qRT-PCR). All estimates are presented over time since first detection of incident malaria; all associations are best described by non-linear models.

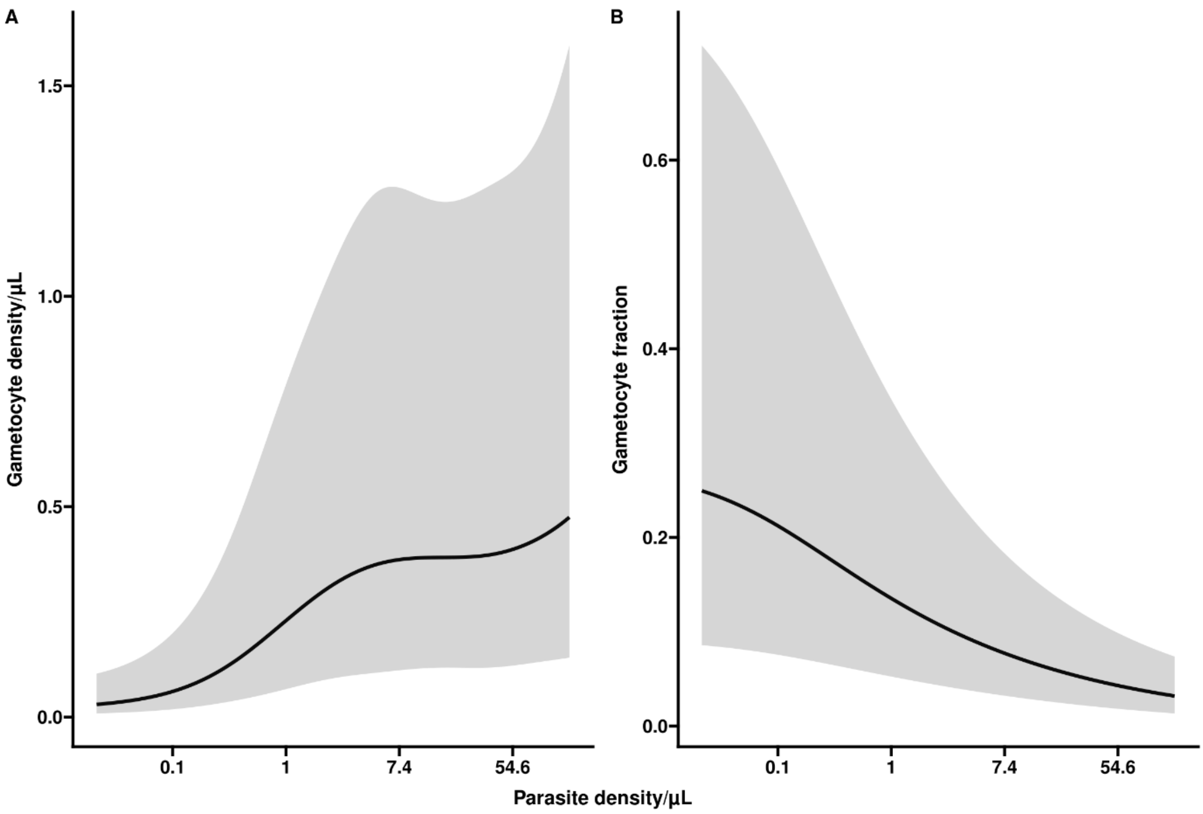


**Figure S6 Gametocyte density and gametocyte fraction in relation to parasite density.**

In panel A, the association of parasite density with gametocyte density is presented, adjusted for the duration of infection. We observe a positive association between total parasite density and gametocyte density. In panel B, the association of parasite density with gametocyte fraction is presented, adjusted for the duration of infection. Higher parasite densities are associated with lower gametocyte fraction at any given moment in time. Visits when the infection was detected without detectable gametocytes were included in these analyses.

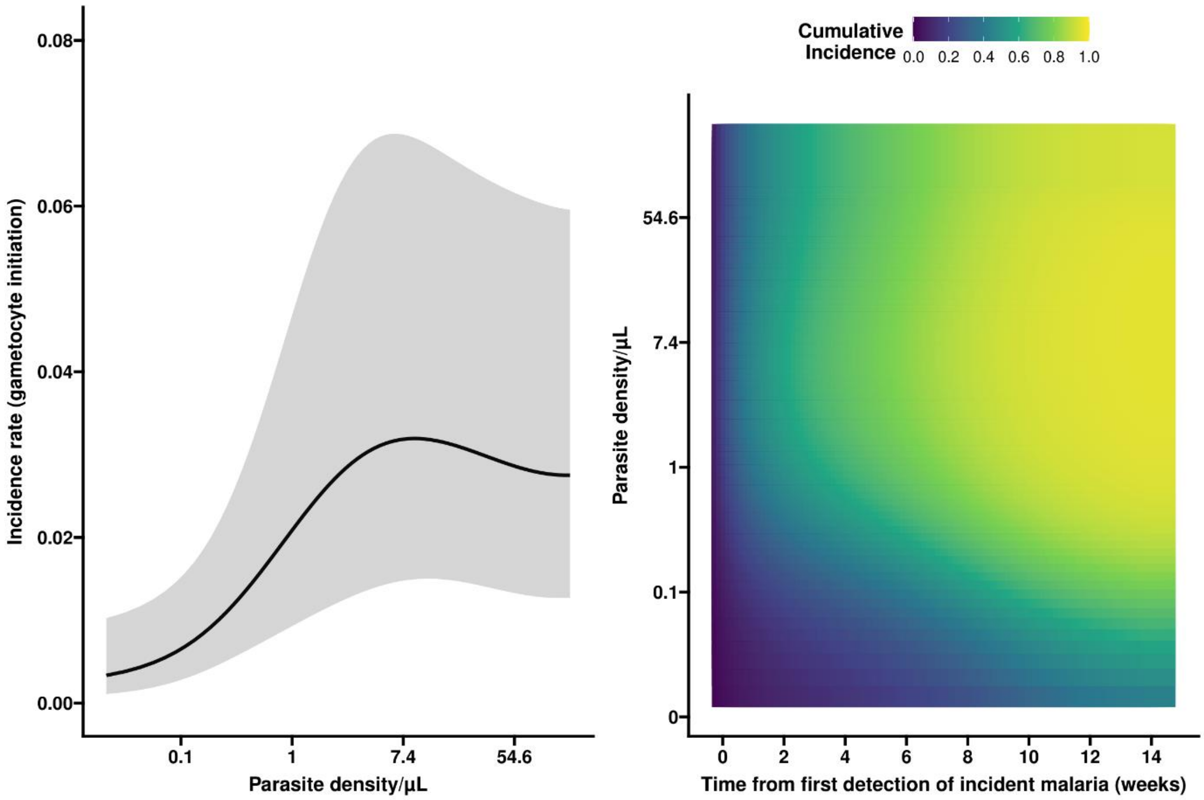


**Figure S7 Gametocyte production in relation to parasite density and infection duration**

Panel A shows the non-linear association between parasite density/µl (log transformed values) and the incidence of gametocyte initiation, adjusted for duration of infection. Panel B presents the association between parasite density and the duration of incident infection with the cumulative incidence of gametocyte initiation. In Figure B, on the y axis the parasite density is expressed per µl, while the x axis describes the time in weeks since the first detection of infection. Low cumulative incidence is shown in blue, whilst cumulative incidences closer to 100% are shown in yellow. Expected cumulative incidences for gametocyte initiation are presented over the full duration of an infection if parasite densities would be maintained at the level given on the y-axis; the likelihood of having initiated gametocyte production increases with increasing parasite density and longer time since infection.

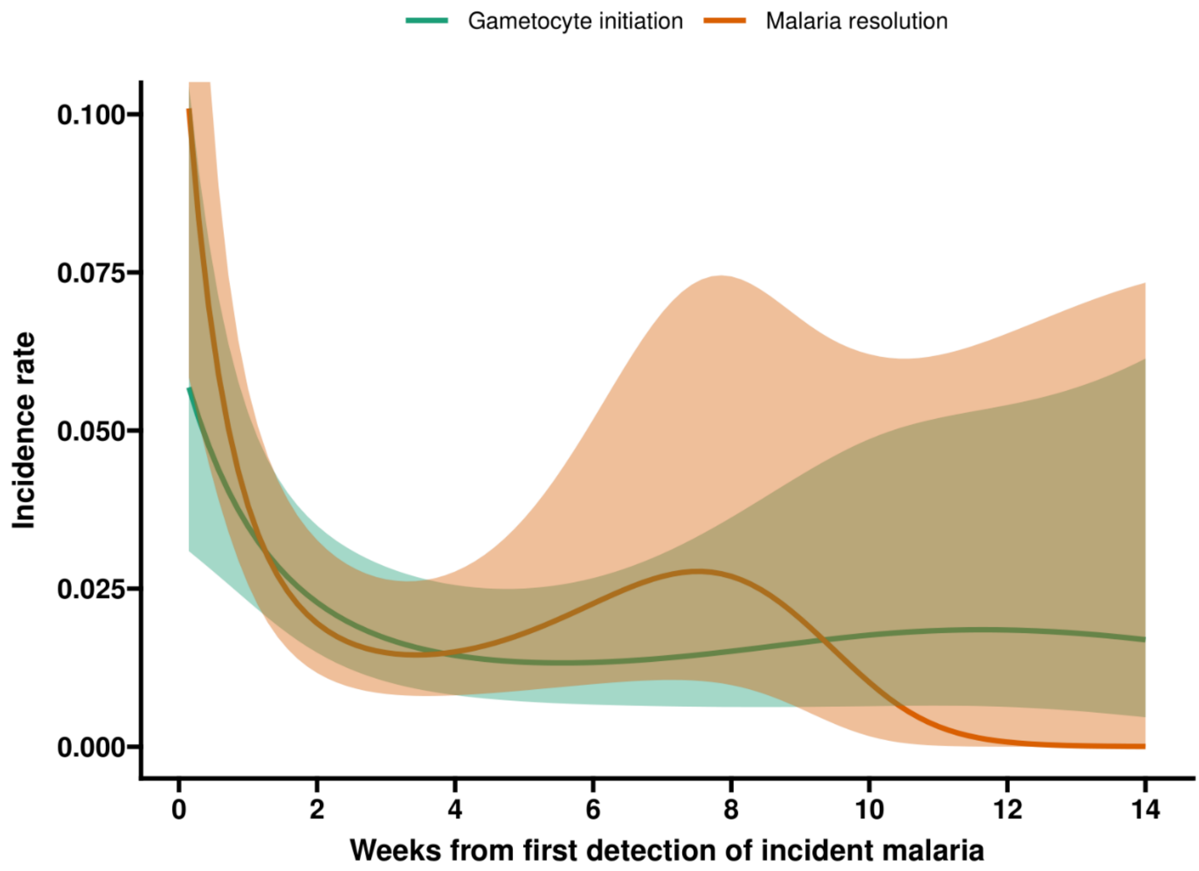


**Figure S8 Baseline incidence density/hazard rates for gametocyte production and resolution without initiating gametocytes over the time since detected incident malaria infection.**

This figure illustrates the unadjusted baseline plots of how the incidence of gametocyte initiation and also malaria resolution prior to gametocyte initiation evolves over time since detected infection.

**Sensitivity analysis for all infections that remained asymptomatic**

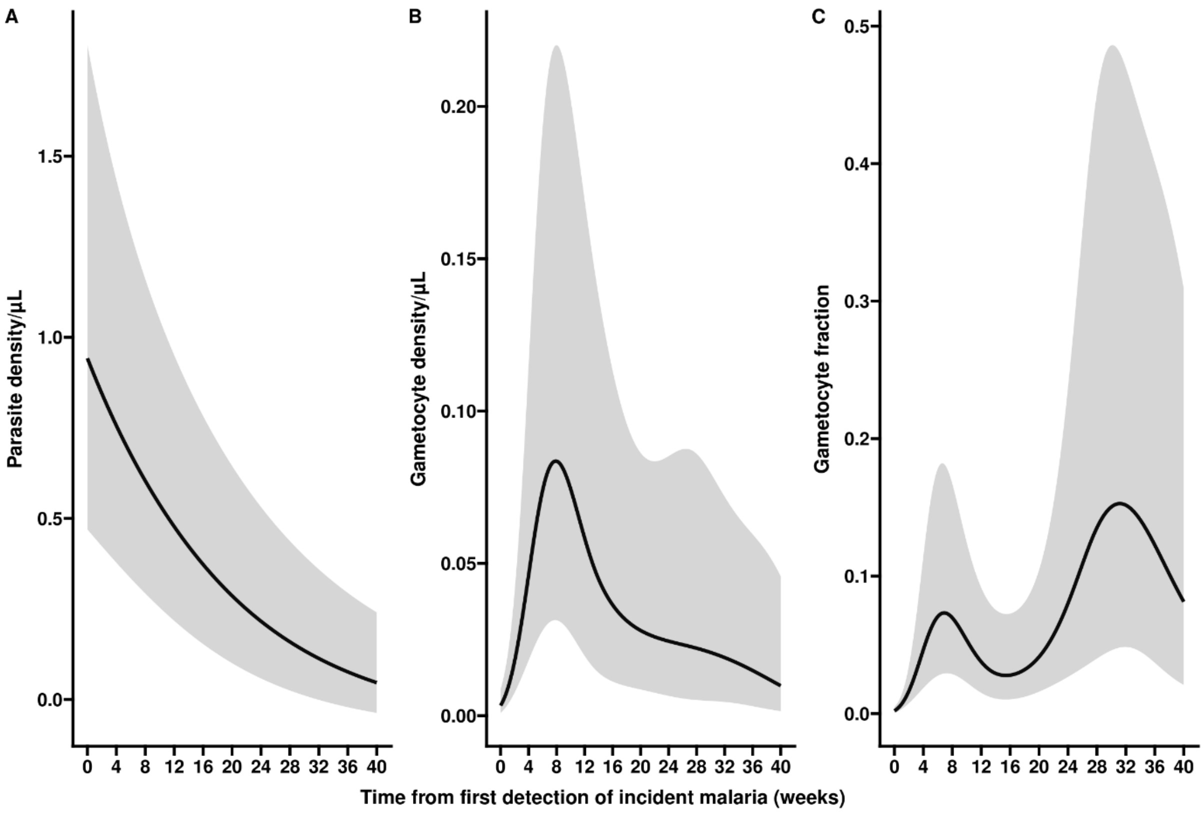


**Figure S9 Parasite density, gametocyte density and gametocyte fraction over the course of incident infections.** Three separate characteristics of infections are presented: parasite density (A), gametocyte density (B) and gametocyte fraction (C). Gametocyte fraction is defined as the proportion of parasites that are gametocytes, estimated as the proportion of the total parasite biomass (i.e. the density estimated by varATS qPCR) that consists of gametocytes (estimated by Ccp4 and PfMGET qRT-PCR). All estimates are presented over time since first detection of incident malaria; all associations are best described by non-linear models.

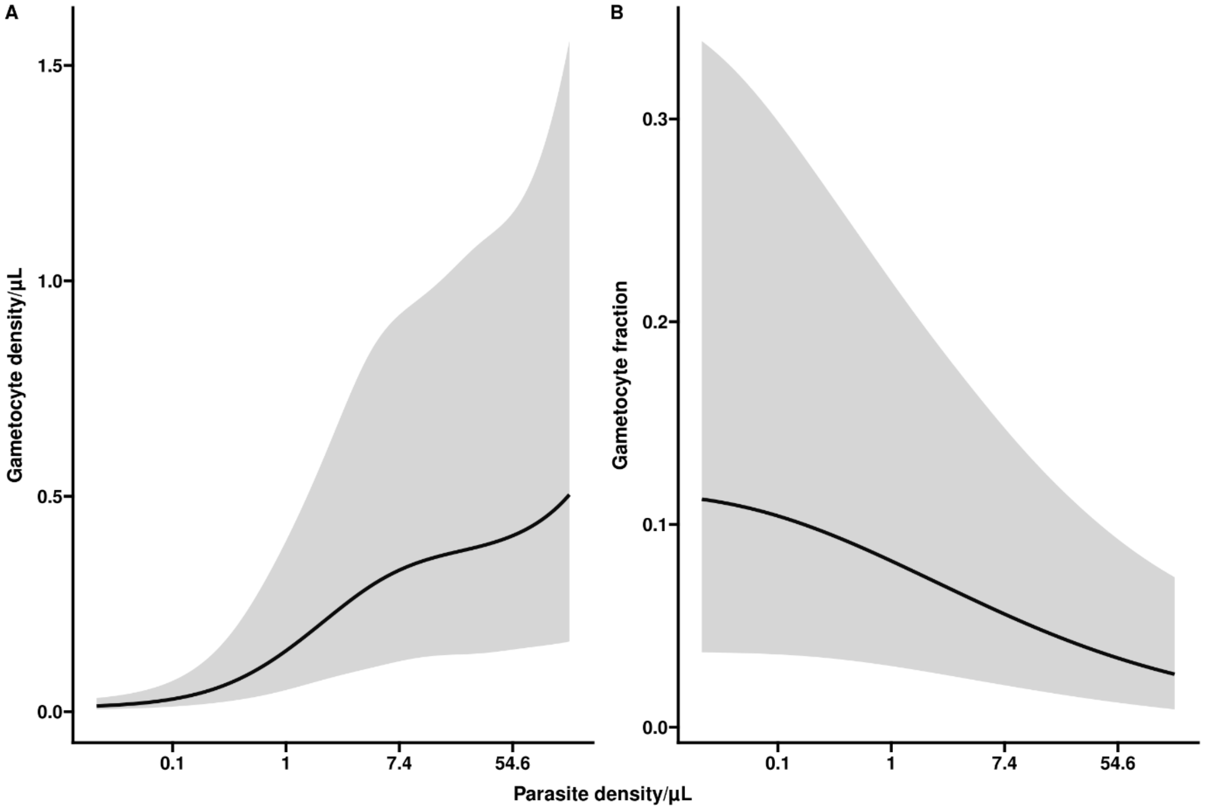


**Figure S10 Gametocyte density and gametocyte fraction in relation to parasite density.** In panel A, the association of parasite density with gametocyte density is presented, adjusted for the duration of infection. We observe a positive association between total parasite density and gametocyte density. In panel B, the association of parasite density with gametocyte fraction is presented, adjusted for the duration of infection. Higher parasite densities are associated with lower gametocyte fraction at any given moment in time. Visits when the infection was detected without detectable gametocytes were included in these analyses.

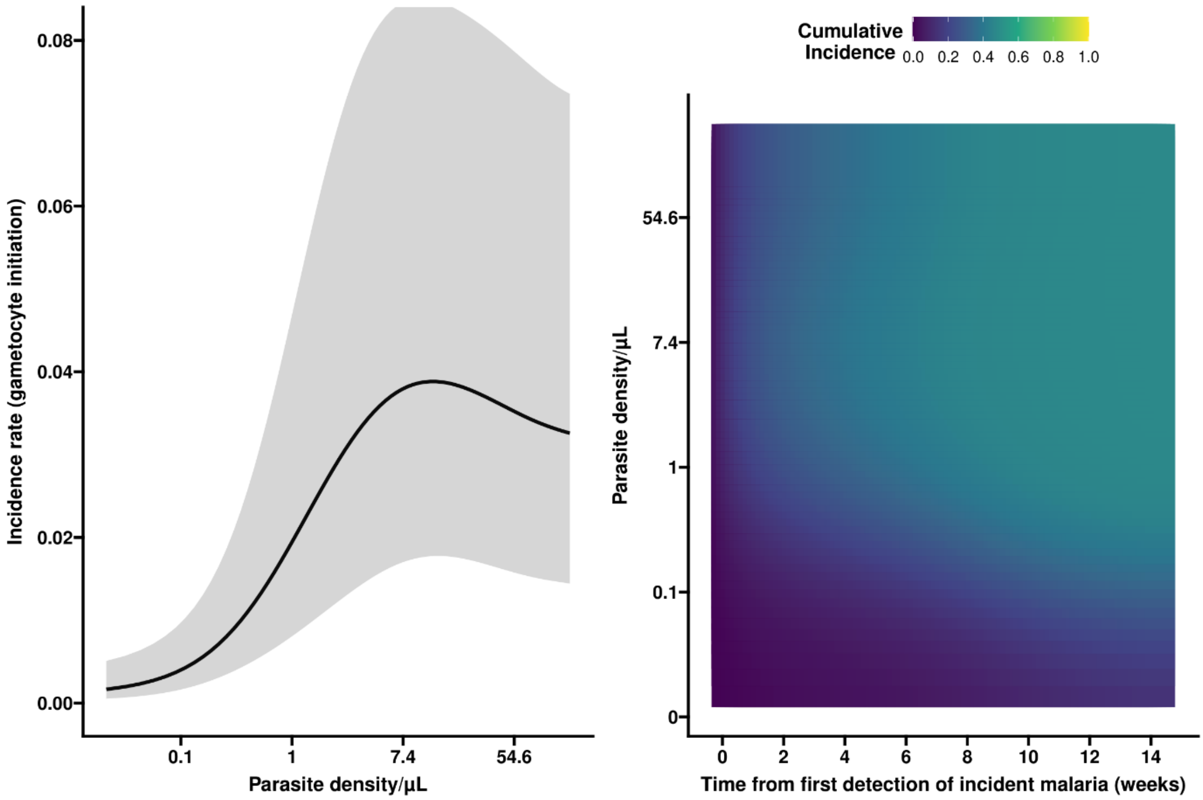


**Figure S11 Gametocyte production in relation to parasite density and infection duration**. Panel A shows the non-linear association between parasite density/µl (log transformed values) and the incidence of gametocyte initiation, adjusted for duration of infection. Panel B presents the association between parasite density and the duration of incident infection with the cumulative incidence of gametocyte initiation. In Figure B, on the y axis the parasite density is expressed per µl, while the x axis describes the time in weeks since the first detection of infection. Low cumulative incidence is shown in blue, whilst cumulative incidences closer to 100% are shown in yellow. Expected cumulative incidences for gametocyte initiation are presented over the full duration of an infection if parasite densities would be maintained at the level given on the y-axis; the likelihood of having initiated gametocyte production increases with increasing parasite density and longer time since infection.

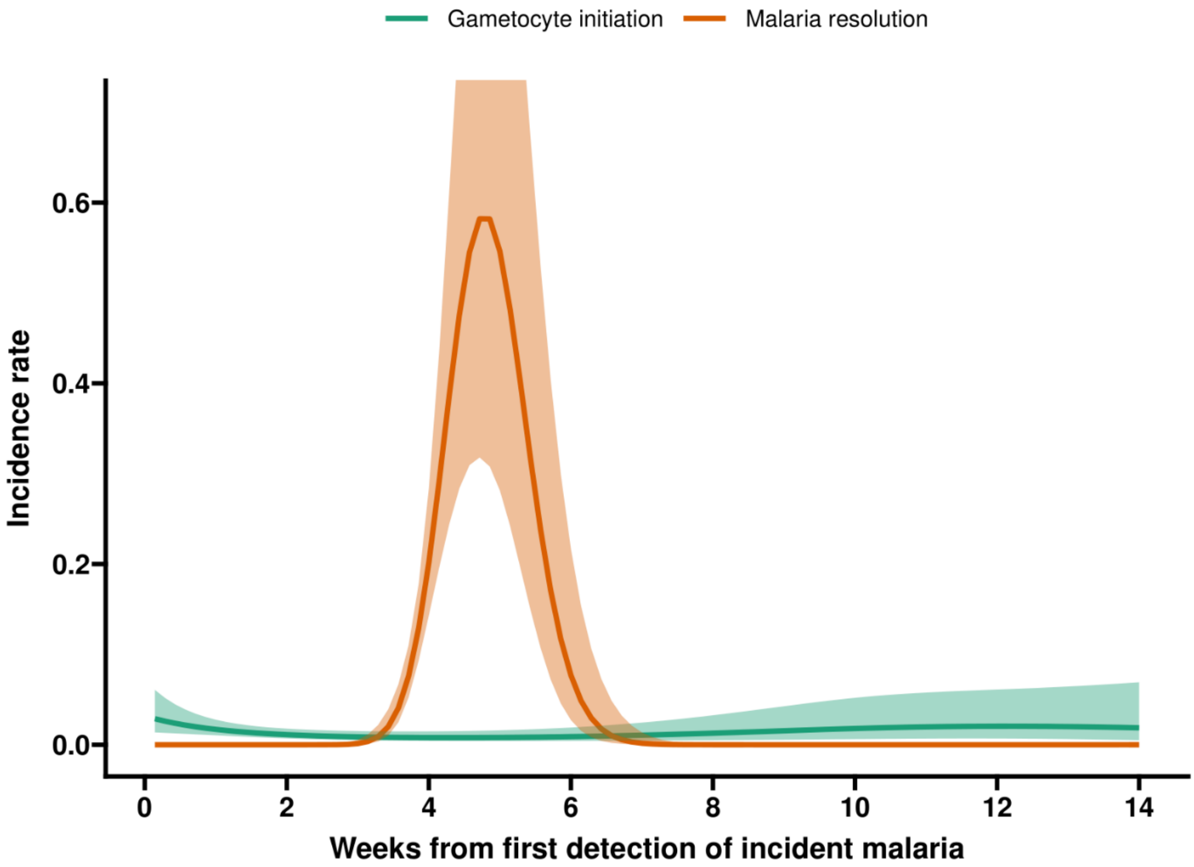


**Figure S12 Baseline incidence density/hazard rates for gametocyte production and resolution without initiating gametocytes over the time since detected incident malaria infection.** This figure illustrates the unadjusted baseline plots of how the incidence of gametocyte initiation and also malaria resolution prior to gametocyte initiation evolves over time since detected infection.

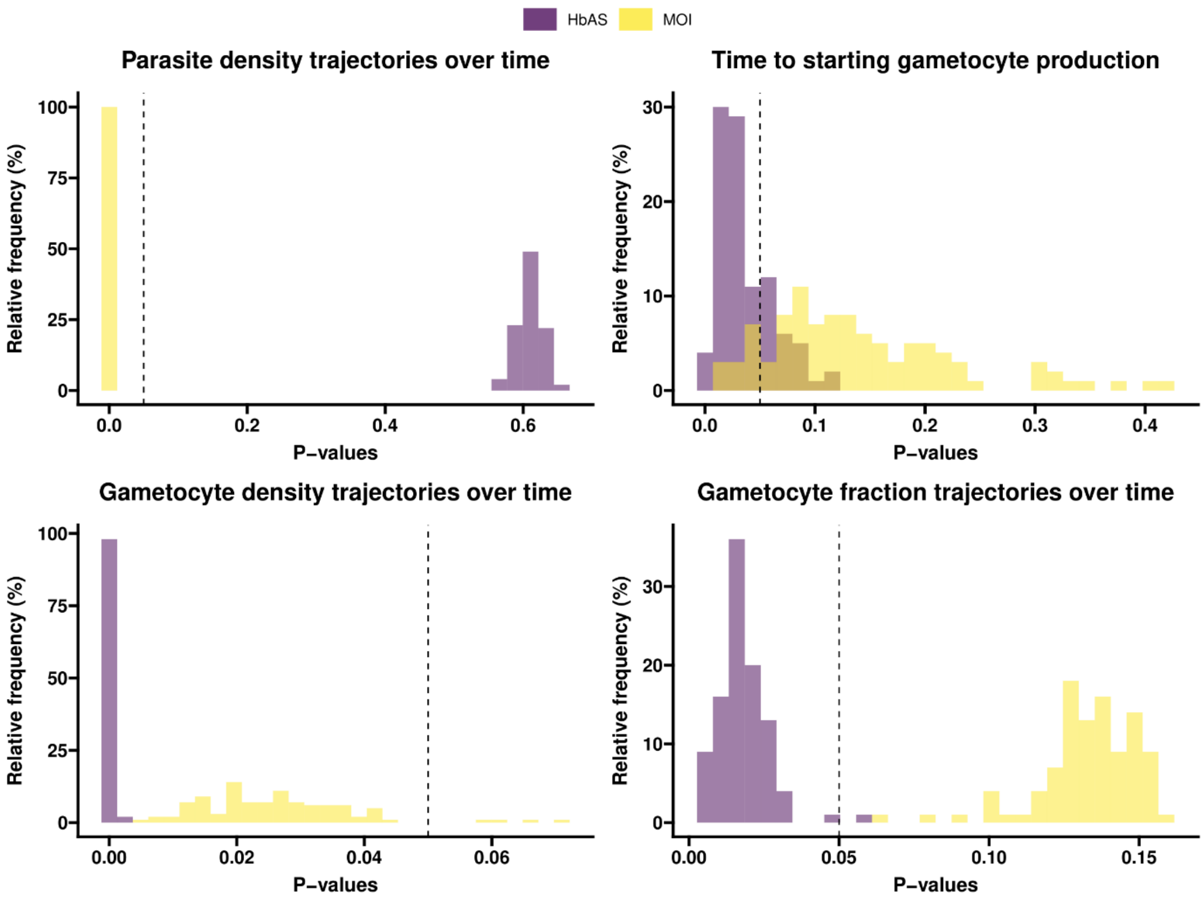


**Figure S13. Distribution of p-values from one hundred imputed datasets to evaluate the sensitivity to HbAS and MOI>1 to interval-censoring.** All underlying exact times, time of incident malaria, time of gametocyte initiation and time of recovery are interval-censored. Ignoring interval-censoring may lead to underestimated standard errors and thus inflated type I error rates. Exact times for each analyses were imputed 100 times from a uniform distribution within the interval, and then each imputed dataset was analyzed for each outcome (**A**) Parasite densities over time, (**B**) time to initiating gametocyte production, (**C**) gametocyte densities over time and (**D**) gametocyte fraction over time) and the p-values for HbAS (purple) and MOI>1 (yellow) were reported. Dashed vertical lines indicated the 5% level of significance cutoff.

| | Parasite density | | Gametocyte incidence | | Gametocyte density | | Gametocyte fraction | | Malaria resolution | |
|---|---|---|---|---|---|---|---|---|---|---|
| | DR(95% CI) | p-value | HR(95% CI) | p-value | DR(95% CI) | p-value | DR(95% CI) | p-value | HR(95% CI) | p-value |
| **<5 years (reference group)** | | | | | | | | | | |
| **5-15 years** | 1.06 (0.11, 10.08) | 0.959 | 1.89 (0.7, 5.1) | 0.1982 | 1.49 (0.27, 8.3) | 0.6423 | 1.19 (0.31, 4.51) | 0.7963 | 0.68 (0.28, 1.63) | 0.375 |
| **16+ years** | 0.14 (0.01, 1.52) | 0.0983 | 1.22 (0.4, 3.74) | 0.7165 | 0.69 (0.11, 4.39) | 0.6879 | 1.06 (0.25, 4.47) | 0.9337 | 0.17 (0.06, 0.47) | 0.0005 |
| **Female (reference group)** | | | | | | | | | | |
| **Male** | 0.46 (0.07, 2.94) | 0.403 | 0.95 (0.44, 2.05) | 0.8855 | 1.18 (0.31, 4.54) | 0.8086 | 1.24 (0.45, 3.42) | 0.6742 | 0.62 (0.27, 1.42) | 0.2505 |
| **AA (reference group)** | | | | | | | | | | |
| **SS** | 106.93 (3.52, 3247.04) | 0.0062 | 1.09 (0.22, 5.38) | 0.9117 | 1.34 (0.11, 16.55) | 0.8137 | 3.55 (0.46, 27.64) | 0.2175 | 1.38 (0.4, 4.85) | 0.6038 |
| **AS** | 1.26 (0.16, 10.18) | 0.8227 | 2.69 (1.14, 6.33) | 0.0209 | 12.55 (2.94, 53.47) | 0.0005 | 3.86 (1.2, 12.36) | 0.0204 | 0.28 (0.09, 0.92) | 0.0326 |
| **MOI=1 (reference group)** | | | | | | | | | | |
| **MOI>1** | 13.62 (2.34, 79.24) | 0.003 | 2.93 (1.15, 7.42) | 0.021 | 5.97 (1.39, 25.6) | 0.014 | 2.37 (0.78, 7.22) | 0.1218 | 0.42 (0.18, 0.97) | 0.0391 |

**Table S1. Sensitivity analysis excluding infections with low parasite densities.** For these analyses, all infections with an initial parasite density below 0.1 parasites per µl were excluded, reducing the study population from 104 monitored infections to 73 infections.

| | Parasite density | | Gametocyte incidence | | Gametocyte density | | Gametocyte fraction | | Malaria resolution | |
|---|---|---|---|---|---|---|---|---|---|---|
| | **DR(95% CI)** | **p-value** | **HR(95% CI)** | **p-value** | **DR(95% CI)** | **p-value** | **DR(95% CI)** | **p-value** | **HR(95% CI)** | **p-value** |
| **<5 years (reference group)** | | | | | | | | | | |
| **5-15 years** | 0.96(0.2, 4.73) | 0.9601 | 2.13 (0.65, 7) | 0.2054 | 1.44 (0.34, 5.98) | 0.6121 | 0.97 (0.22, 4.25) | 0.9725 | 1.18 (0.55, 2.53) | 0.6736 |
| **16+ years** | 1 (0.2, 4.95) | 0.9963 | 2.02 (0.56, 7.31) | 0.2727 | 1.03 (0.25, 4.34) | 0.9625 | 1.42 (0.32, 6.22) | 0.6359 | 0.8 (0.35, 1.82) | 0.5834 |
| **Female (reference group)** | | | | | | | | | | |
| **Male** | 1.24 (0.38, 4.06) | 0.7151 | 1.25 (0.51, 3.07) | 0.6172 | 1.71 (0.61, 4.82) | 0.3 | 1.43 (0.5, 4.11) | 0.5013 | 0.98 (0.51, 1.86) | 0.9486 |
| **AA (reference group)** | | | | | | | | | | |
| **SS** | 0.13 (0.01, 2.06) | 0.1409 | 0 (0, Inf) | 1 | 1.86 (0.1, 35) | 0.671 | 0.26 (0.01, 5.22) | 0.3695 | 0.89 (0.22, 3.56) | 0.8701 |
| **AS** | 3.13 (0.81, 12.06) | 0.091 | 3.02 (1.18, 7.74) | 0.0188 | 8.45 (2.77, 25.82) | 0.0001 | 4.31 (1.29, 14.37) | 0.0152 | 1.16 (0.5, 2.67) | 0.7224 |
| **MOI=1 (reference group)** | | | | | | | | | | |
| **MOI>1** | 11.47 (2.8, 47.02) | 0.0005 | 2.75 (0.77, 9.74) | 0.1103 | 4.46 (0.83, 24.06) | 0.0759 | 3.07 (0.54, 17.38) | 0.195 | 1.03 (0.15, 7.06) | 0.9729 |

**Table S2. Sensitivity analysis of asymptomatic infections.** In these analyses all infections that were either symptomatic at the moment of infection detection or at any time-point during follow-up were removed from the analyses. This reduced the study population from 104 monitored infections to 76 infections.
